# Clinical course and outcomes of critically ill COVID-19 patients in two successive pandemic waves

**DOI:** 10.1101/2021.02.26.21251848

**Authors:** Athanasios Chalkias, Ioannis Pantazopoulos, Nikolaos Papagiannakis, Anargyros Skoulakis, Eleni Laou, Konstantina Kolonia, Nicoletta Ntalarizou, Dimitrios Ragias, Christos Kampolis, Luis García de Guadiana Romualdo, Konstantinos Tourlakopoulos, Athanasios Pagonis, Salim S Hayek, Jesper Eugen-Olsen, Konstantinos Gourgoulianis, Eleni Arnaoutoglou

## Abstract

**Rationale:** The progress of COVID-19 from moderate to severe may be precipitous, while the heterogenous characteristics of the disease pose challenges to the management of these patients.

**Objectives:** To characterize the clinical course and outcomes of critically ill patients with COVID-19 during two successive waves.

**Methods:** We leveraged the multi-center SuPAR in Adult Patients With COVID-19 (SPARCOL) study and collected data from consecutive patients requiring admission to the intensive care unit from April 1^st^ to December 31^st^, 2020.

**Measurements and Main Results:** Of 252 patients, 81 (32%) required intubation and mechanical ventilation. Of them, 17 (20.9%) were intubated during the first wave, while 64 (79%) during the second wave. The most prominent difference between the two waves was the overall survival (first wave 58.9% *vs.* second wave 15.6%, adjusted p-value=0.006). This difference is reflected in the prolonged hospitalization during the first wave. The mean ICU length of stay (19.1 *vs.* 11.7 days, p=0.022), hospital length of stay (28.5 *vs.* 17.1 days, p=0.012), and days on ventilator (16.7 *vs.* 11.5, p=0.13) were higher during the first wave. A significant difference between the two waves was the development of bradycardia. In the first wave, 2 (11.7%) patients developed sinus bradycardia only after admission to the intensive care unit, while in the second wave, 63 (98.4%) patients developed sinus bradycardia during hospitalization.

**Conclusions:** Survival of critically ill patients with COVID-19 was significantly lower during the second wave. The majority of these patients developed sinus bradycardia during hospitalization.

## Introduction

Although several months have been passed after the inception of the SARS-CoV-2 pandemic, the global numbers of critically ill patients with severe coronavirus disease 2019 (COVID-19) are increasing in many European countries. The progress of the disease from moderate to severe may be precipitous requiring life-sustaining interventions and admission to the intensive care unit (ICU) (1,2). For example, in the early stages of the disease, patients may be characterized by mild hypoxemia and a hyperdynamic circulatory state with high cardiac index (3), but as the disease progresses, acute respiratory distress syndrome (ARDS), high left ventricular filling pressure, and heart failure may ensue (4,5).

The heterogenous characteristics of COVID-19 pose challenges to the management of these patients (6). Intubation and mechanical ventilation may improve outcome, but may aggravate lung injury and induce circulatory derangement as well. Of note, the 24-h mortality after tracheal intubation has been reported 2-10.4% (7,8), but the effects of peri-intubation interventions are largely unknown and may differ between outbreaks. In addition, a range of multiorgan complications following COVID-19 infection may develop and further aggravate the clinical course and prognosis (9).

To better describe the clinical course and outcomes of critically ill patients with COVID-19, we leveraged the multi-center SuPAR in Adult Patients With COVID-19 (SPARCOL) study to assess the clinical characteristics and interventions used in patients requiring ICU admission during the two COVID-19 spikes.

## Materials and methods

### The SuPAR in Adult Patients With COVID-19 study

The SuPAR in Adult Patients With COVID-19 is an ongoing multi-center observational study (ClinicalTrials.gov Identifier: NCT04590794) which primary purpose is to characterize levels of biomarker soluble urokinase plasminogen activator receptor (suPAR) and its association with respiratory complications, admission to ICU, organ injury, and survival of patients with COVID-19. Participating centers include: University of Thessaly, Larisa, Greece; the Hippokration University Hospital, Athens, Greece; the Evangelismos University Hospital, Athens, Greece; and the University of Copenhagen at Hvidovre, Denmark. Ethical approval was provided by the Ethical Committee of the University Hospital of Larisa (IRB no. 17543), Larisa, Greece on 24 April 2020. The study was performed according to national and international guidelines. Written informed consent was obtained from the patients.

Inclusion criteria for this study were: (1) adult (≥18 years old) patients hospitalized primarily for COVID-19; (2) a confirmed SARS-CoV-2 infection diagnosed through reverse transcriptase polymerase chain reaction test of nasopharyngeal or oropharyngeal samples; (3) at least one blood sample collected at admission and stored for biomarker testing; and (4) admission to ICU.

### Study design and outcomes definitions

For the purpose of this study, we collected data from consecutive patients hospitalized for COVID-19 (n=252) during the period of April 1^st^ to December 31^st^, 2020, the date the database was locked for the purpose of this analysis. Furthermore, we divided the study period into two smaller ones according to the duration of the two COVID-19 waves in most European countries (March 2020 - July 2020 and August 2020 - December 2020).

Local investigators screened and reported all intubations occurring in the emergency department, ICU, and wards during the study period. Manual chart review was used to gather details of the demographics and past medical history, peri-intubation period, laboratory studies, ICU course, and outcomes. All patients were followed until the 30^th^ day post-discharge or death. We excluded from this analysis patients with confirmed SARS-CoV-2 infection who were not primarily admitted for COVID-19, patients with incomplete data, patients with pre-existing severe cardiac or respiratory disease, such as heart failure, more than mild chronic obstructive pulmonary disease, or pulmonary vascular disease, patients with pacemaker or implantable cardioverter-defibrillator, and patients who underwent intubation following a cardiac arrest.

The primary aim of the study was to assess the differences in the clinical characteristics and outcome of critically ill patients with COVID-19 requiring ICU admission between the two waves. Secondary aims were to evaluate the incidence and nature of major complications during hospitalization, including intubation-related major complications, and characterize levels of various inflammatory biomarkers and their association with outcomes of patients with COVID-19. Intubation-related complications were defined as the occurrence of at least one of the following events: (a) aggravation of hypoxemia (defined as SpO_2_<80% or by a decrease in the pre-induction SpO_2_ value of more than 20%); (b) severe cardiovascular collapse [systolic arterial pressure (SAP)<65 mmHg recorded once or SAP <90 mmHg for >30 minutes or new need/increase of vasopressor/inotrope support and/or fluid load > 15 ml/kg); (c) cardiac arrhythmia; (d) cardiac arrest; (e) other outcomes (incidence of difficult intubation, ‘cannot intubate cannot oxygenate’ scenario, emergency front of neck airway, esophageal intubation, dental injury or airway injury, pneumothorax, pneumomediastinum, subcutaneous emphysema, or aspiration of gastric contents).

We followed the Strengthening the Reporting of Observational Studies in Epidemiology (STROBE) statement guidelines for observational cohort studies (10).

### Data collection and monitoring

The data collected included details at presentation, past medical history, home medications, hospitalization course, and outcomes. Laboratory testing included general blood count; biochemical profile including levels of BUN, creatinine, protein, albumin, high-sensitive C-reactive protein (hs-CRP), ferritin, D-Dimer, and lactate dehydrogenase; arterial blood gases; and suPAR levels at admission. All serum samples were obtained on admission to hospital, before any treatment or non-invasive/invasive ventilation. The data analysis was based on predefined data points on a prospective data collection form. The authors and laboratory technicians were blinded to clinical data and measurements until the end of the study and all data were analyzed. Also, an independent data and safety monitoring research staff monitored safety, ethical, and scientific aspects of the study, while an independent enrollment research staff was responsible for exclusion of all patients not meeting inclusion criteria.

### Statistical analysis

Statistical analysis was performed using R v4.0. The non-parametric Mann-Whitney test was used to observe differences between numerical observations in the first and second wave. Additionally, the chi-square test of independence was applied to the categorical observations. In both cases, the Benjamini–Hochberg false discovery rate correction was applied in the resulting P values to account for the multiple number of tests. Adjusted p-values less than 0.05 were deemed significant. Spearman’s rho coefficient was used for linear correlation. To assess the impact of the variables of interest (summarized in Table 5) to overall survival, we have fitted a logistic regression statistical model with survival as the dependent variable and the other variables as independent variables. For each variable the respective odds ratio and p-value was computed. No adjustment was applied in the resulting p-values from this model.

## Results

During the period April 1^st^ to December 31^st^, 252 consecutive patients were hospitalized for COVID-19 and 81 (32%) required intubation and mechanical ventilation and were included in the study. Of them, 17 (20.9%) were intubated during the first wave, while 64 (79%) during the second wave. Their demographic and clinical characteristics are presented in Tables 1 and 2.

**Table 1.** Demographics and clinical characteristics at presentation

|  | First wave (n=17) | Second wave (n=64) | Adjusted p-value |
| --- | --- | --- | --- |
| **Demographics** |  |  |  |
| Sex |  |  |  |
| Female, n (%) | 8 (47%) | 20 (31%) | 0.747 |
| Male, n (%) | 9 (53%) | 44 (69%) |  |
| Age (years), mean (95% CI) | 65.23 (58.26 - 72.21) | 68.67 (65.83 - 71.52) | 0.457 |
| Height (cm), mean (95% CI) | 169.29 (164.96 - 173.63) | 171.65 (169.98 - 173.34) | 0.346 |
| Weight (kg), mean (95% CI) | 85.33 (57.97 - 112.7) | 84.56 (79.88 - 89.25) | 0.913 |
| Body mass index (kg/m<sup>2</sup>), mean (95% CI) | 28.74 (24.39 - 33.1) | 27.57 (26.6 - 28.56) | 1 |
| **Presentation** |  |  |  |
| *Vital signs* |  |  |  |
| Systolic arterial pressure (mmHg), mean (95% CI) | 135.05 (128.98 - 141.14) | 131.79 (127.29 - 136.31) | 0.625 |
| Diastolic arterial pressure (mmHg), mean (95% CI) | 64.35 (59.06 - 69.65) | 73.14 (69.95 - 76.33) | 0.055 |
| Mean arterial pressure (mmHg), mean (95% CI) | 87.93 (82.92 - 92.96) | 92.75 (89.4 - 96.1) | 0.347 |
| Pulse pressure (mmHg), mean (95% CI) | 69.23 (63.12 - 75.35) | 58.71 (55.36 - 62.08 ) | 0.023 |
| Heart rate (min<sup>-1</sup>), mean (95% CI) | 80.23 (70.92 - 89.55) | 82 (77.19 - 86.81) | 0.861 |
| Respiratory rate (min<sup>-1</sup>), mean (95% CI) | 23.41 (21.58 - 25.24) | 26.34 (25.53 - 27.16) | **0.027** |
| SpO<sub>2</sub> (%), mean (95% CI) | 84.82 (83.56 - 86.09) | 83.51 (82.6 - 84.43) | 0.306 |
| Capillary refill time (s), mean (95% CI) | 3.17 (2.57 - 3.79) | 3.26 (2.87 - 3.66 ) | 0.998 |
| Temperature (°C), mean (95% CI) | 38.36 (37.94 - 38.79) | 38.66 (38.51 - 38.82) | 0.347 |
| *Biomarkers, blood tests, and radiographic findings* |  |  |  |
| suPAR (ng/mL), mean (95% CI) | 7.32 (4.64 - 10.02) | 9.29 (8.16 - 10.44) | 0.105 |
| White blood cells (K/uL), mean (95% CI) | 5.4 (3.85 - 6.95) | 7.75 (6.71 - 8.8 ) | 0.105 |
| Hemoglobin (g/dL), mean (95% CI) | 13.27 (12.56 - 13.98) | 12.3 (11.85 - 12.75) | 0.141 |
| Platelets (K/uL), mean (95% CI) | 194.17 (158.61 - 229.74) | 219.9 (196.81 - 243.01) | 0.463 |
| BUN (mg/dL), mean (95% CI) | 39.3 (30.77 - 47.83) | 61.62 (53.1 - 70.15) | 0.053 |
| Creatinine (mg/dL), mean (95% CI) | 1 (0.83 - 1.17) | 1.3 (1.07 - 1.54) | 0.831 |
| Protein (g/dL), mean (95% CI) | 6.7 (6.37 - 7.04) | 6.26 (6.09 - 6.44) | 0.088 |
| Albumin (g/dL), mean (95% CI) | 4.09 (3.86 - 4.33) | 3.46 (3.37 - 3.57 ) | **<0.001** |
| hs-CRP (mg/dL), mean (95% CI) | 6.07 (2.99 - 9.17) | 12.02 (9.24 - 14.81) | 0.111 |
| Ferritin (ng/mL), mean (95% CI) | 1335.77 (388.27 - 2283.28) | 1079 (866.69 - 1291.41) | 0.744 |
| D-Dimer (ng/mL), mean (95% CI) | 733.47 (296.67 - 1170.27) | 952.4 (659.04 - 1245.77) | 0.351 |
| Lactate dehydrogenase (IU/L), mean (95% CI) | 375.47 (210.97 - 539.97) | 442.93 (383.86 - 502.01) | 0.201 |
| Chest radiography |  |  |  |
| Bilateral lung opacities, n (%) | 15 (88%) | 55 (86%) | 0.997 |
| Pleural effusion, Bilateral lung opacities, n (%) | 2 (12%) | 4 (6%) |  |
| Pleural effusion, Bilateral lung opacities, Pulmonary contusion, n (%) | 0 (0%) | 2 (3%) |  |
| Pleural effusion, Bilateral lung opacities, Pulmonary embolism, n (%) | 0 (0%) | 1 (2%) |  |
| Pulmonary contusion, n (%) | 0 (0%) | 2 (3%) |  |
| *Severity scores* |  |  |  |
| APACHE II, mean (95% CI) | 6.64 (5.78 - 7.52) | 6.46 (6.04 - 6.9) | 0.814 |
| SOFA, mean (95% CI) | 2.52 (2.21 - 2.85) | 2.64 (2.38 - 2.9 ) | 0.833 |

SpO<sub>2</sub>, peripheral capillary oxygen saturation; BUN, Blood urea nitrogen; hs-CRP, high-sensitivity C-reactive protein

**Table 2.**
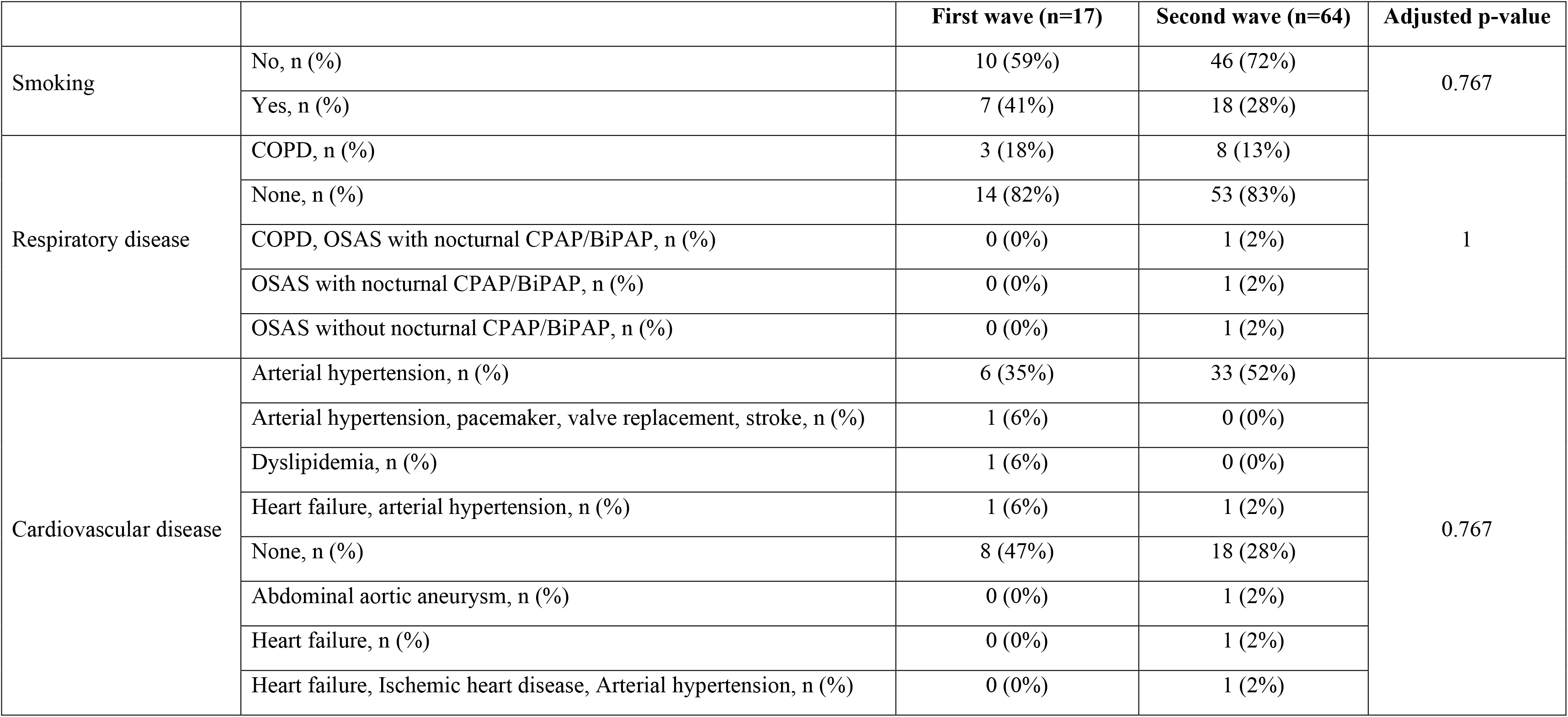
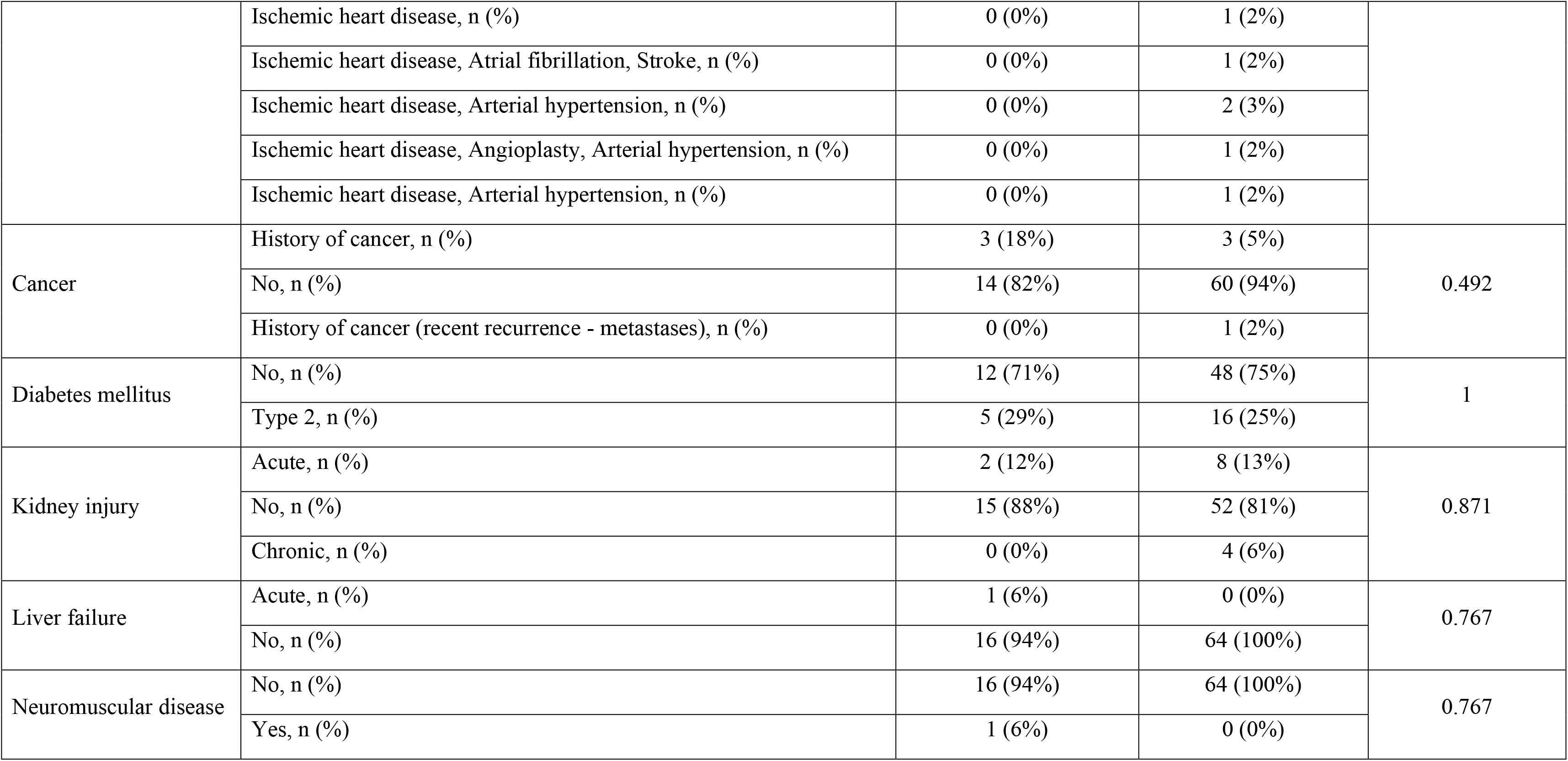
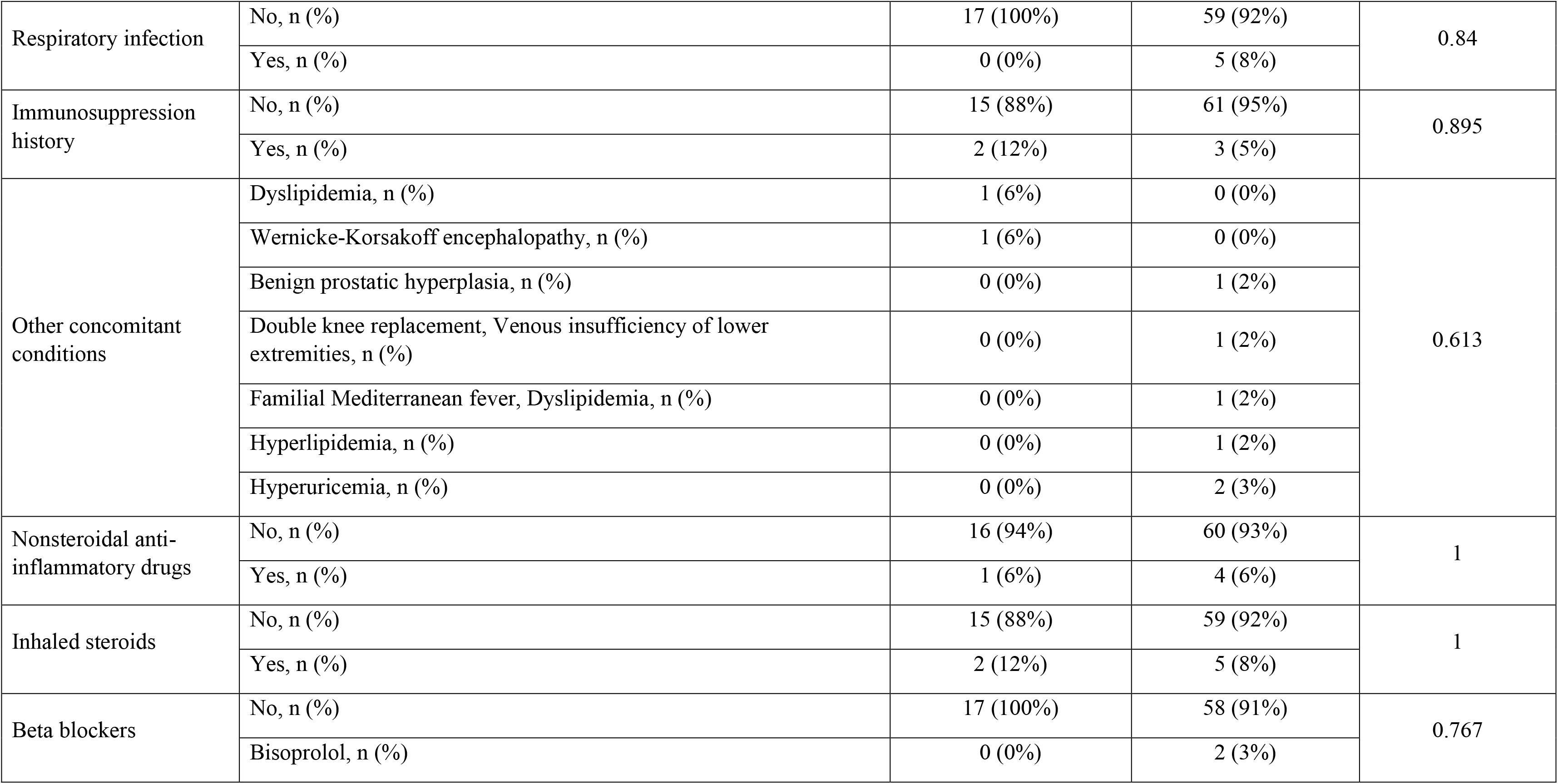
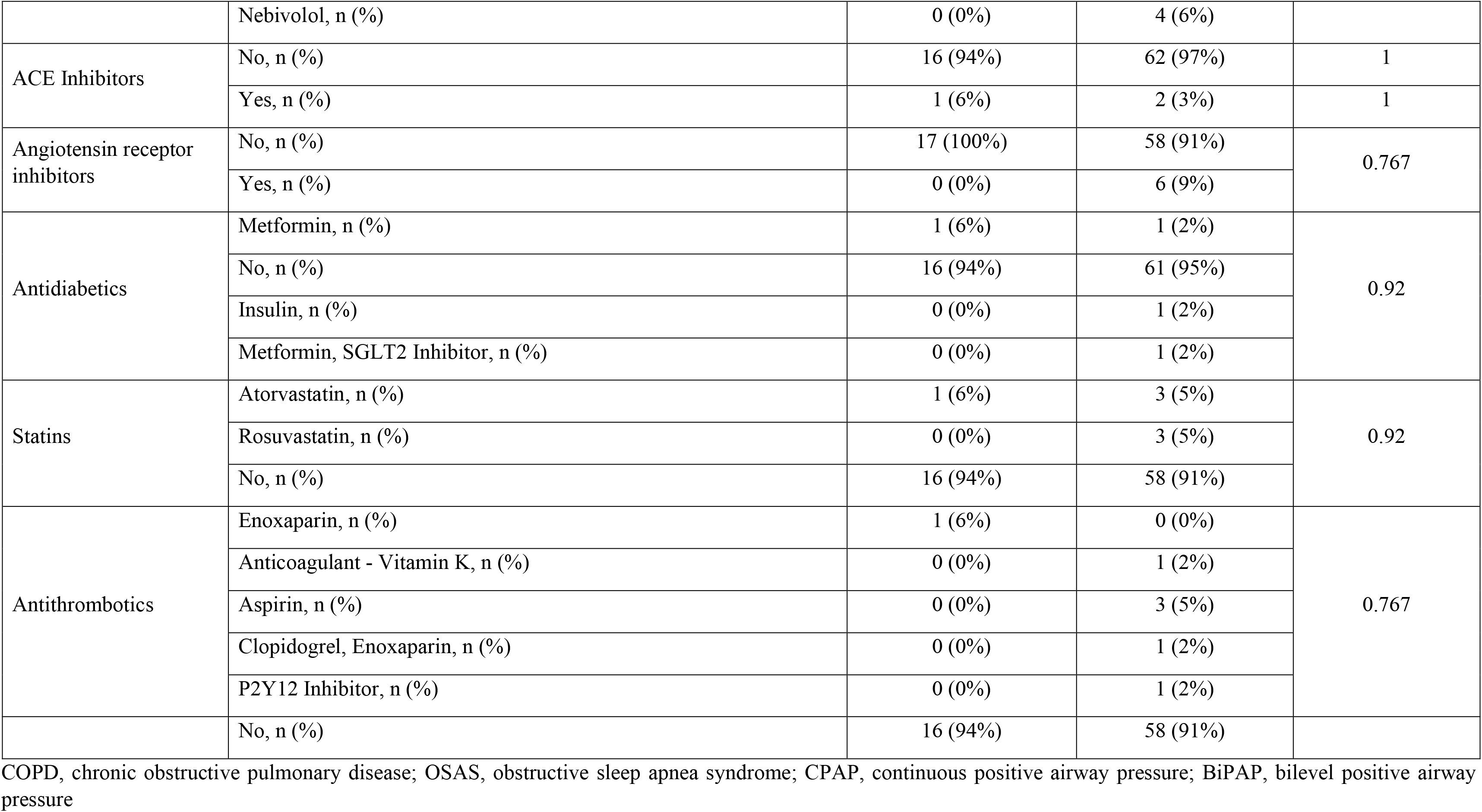
Medical history of study population

A significant difference between the two waves was the development of bradycardia after admission. In the first wave, 2 (11.7%) patients developed sinus bradycardia only after ICU admission (at the 14^th^ and 28^th^ day, respectively), while in the second wave, 63 (98.4%) patients developed sinus bradycardia before admission to ICU (Fig. 1). In the second wave, the average time for development of bradycardia was 5 and 10 days from admission and onset of symptoms, respectively. In all patients, bradycardia developed without clinically detectable myocardial necrosis, while none of them was receiving drugs inducing bradycardia, such as hydroxychloroquine, moxifloxacin, azithromycin, or remdesivir. In all patients, bradycardia was diagnosed and monitored with serial electrocardiograms, cardiac telemetry, and/or continuous electrocardiography.

**Figure 1.**
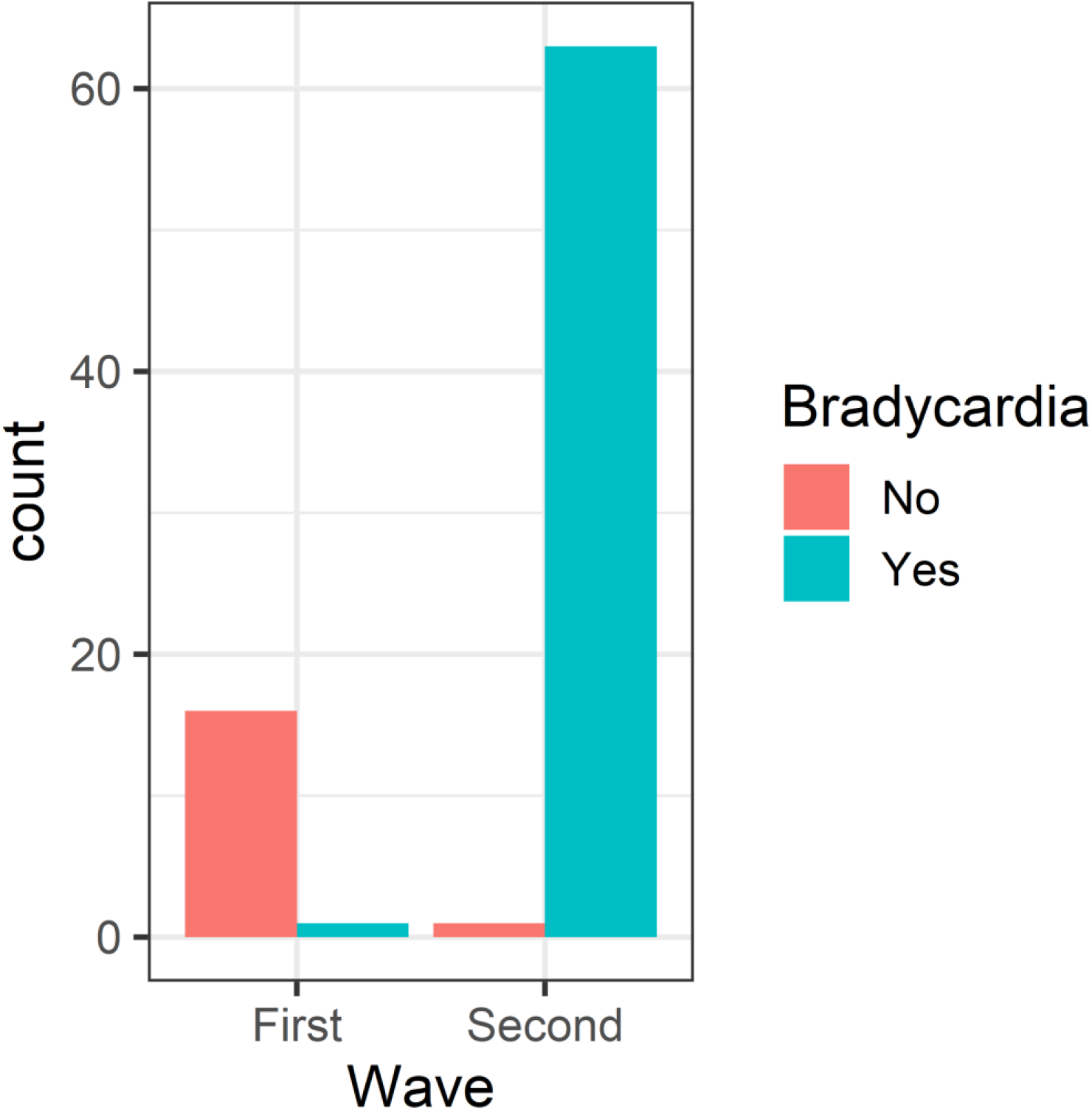
The incidence of bradycardia during the two waves.

### Peri-intubation period

Oxygen administration during laryngoscopy was used in both waves, with high-flow nasal cannula being the preferred method during the second one (p<0.001). However, oxygen administration during laryngoscopy did not seem to prevent desaturation during or after intubation (p=0.53). Peri-intubation characteristics and complications are presented in Table 3, as well as in Tables E1 and E2 in the online data supplement. No difference in overall survival (p=0.85) and risk of developing any post-intubation complications (p=0.28) were found in patients intubated using rapid *vs.* delayed sequence intubation.

**Table 3.** Post-intubation complications (within 30 minutes after endotracheal tube placement confirmation)

|  |  | First wave (n=17) | Second wave (n=64) | Adjusted p-value |
| --- | --- | --- | --- | --- |
| Need of new/modification of vasopressor/inotrope support after intubation | No, n (%) | 16 (94%) | 45 (70%) | 0.54 |
|  | Yes, n (%) | 1 (6%) | 19 (30%) |  |
| Severe cardiovascular collapse | No, n (%) | 0 (0%) | 18 (28%) | 0.157 |
|  | Yes, n (%) | 17 (100%) | 46 (72%) |  |
| Aspiration of gastric contents | No, n (%) | 16 (94%) | 62 (97%) | 0.993 |
|  | Yes, n (%) | 1 (6%) | 2 (3%) |  |
| Dental injury | No, n (%) | 15 (88%) | 63 (98%) | 1 |
|  | Yes, n (%) | 2 (12%) | 1 (2%) |  |
| Pneumothorax and/or pneumomediastinum and/or subcutaneous emphysema | No, n (%) | 17 (100%) | 60 (94%) | 0.823 |
|  | Yes, n (%) | 0 (0%) | 4 (6%) |  |
| Cardiac arrest | Death, n (%) | 1 (6%) | 2 (3%) | 0.997 |
|  | No, n (%) | 16 (0%) | 61 (95%) |  |
|  | With return of spontaneous circulation, n (%) | 0 (0%) | 1 (2%) |  |

AV, atrioventricular

### Clinical course and outcome after admission to ICU

The most prominent difference between the two waves was the overall survival (first wave 58.9% *vs.* second wave 15.6%, adjusted p-value=0.006). This difference is reflected in the prolonged hospitalization during the first wave. The mean ICU length of stay during the first and second wave was 19.1 and 11.7 days (p=0.022), respectively. Accordingly, the mean hospital length of stay was 28.5 and 17.1 days (p=0.012), respectively, while the number of days on ventilator were also greater in the first wave (16.7 *vs.* 11.5, p=0.13) (Table 4). In multivariable analysis, only increasing age was associated with mortality (Table 5).

**Table 4.** Main outcomes of the study

|  |  | First wave (N=17) | Second wave (N=64) | Adjusted p-value |
| --- | --- | --- | --- | --- |
| Days on ventilator, (days), mean (95% CI) |  | 16.706 (10.81 - 22.6) | 11.516 (9.81 - 13.22) | 0.13 |
| ICU Length of stay, (days), mean (95% CI) |  | 19.176 (13.64 - 24.71) | 11.719 (10.02 - 13.42) | **0.022** |
| Hospital length of stay, (days), mean (95% CI) |  | 28.588 (21.63 - 35.55) | 17.079 (15.21 - 18.95) | **0.012** |
| Reintubation (reason) | No, n (%) | 17 | 63 | 1 |
|  | Acute coronary syndrome, n (%) | 0 | 1 |  |
| Final outcome | Death, n (%) | 7 | 54 | **0.006** |
|  | Discharge, n (%) | 10 | 10 |  |
| Cause of death | Primary cardiovascular etiology, n (%) | 1 | 4 | **0.012** |
|  | Respiratory failure or MODS, n (%) | 3 | 42 |  |
|  | Respiratory failure or MODS, Sepsis, n (%) | 3 | 6 |  |
|  | Primary cardiovascular etiology, Sepsis, n (%) | 0 | 2 |  |
| Readmission | No, n (%) | 10 | 9 | 1 |
|  | Yes, n (%) | 0 | 1 |  |

ICU, intensive care unit, MODS, Multiple Organ Dysfunction Syndrome

**Table 5.** Multivariable analysis of the association between patient characteristics and survival

|  | Survival |  |
| --- | --- | --- |
|  | Odds Ratio (Std Error) | p-value |
| Age (per year increase) | 0.84 (0.06) | **0.004** |
| Sex | 0.442 (0.81) | 0.313 |
| Body mass index | 0.849 (0.08) | 0.052 |
| Cardiovascular | 1.434 (0.99) | 0.717 |
| Diabetes mellitus | 2.391 (0.88) | 0.321 |
| Statin use | 8.027 (1.47) | 0.158 |
| P/F Ratio pre-intubation | 0.932 (0.07) | 0.317 |
| Creatinine (per mg/dL increase) | 1.829 (0.59) | 0.309 |
| Systolic arterial pressure (per mmHg increase) | 1.67 (0.86) | 0.55 |
| Diastolic arterial pressure (per mmHg increase) | 2.568 (1.95) | 0.628 |
| Mean arterial pressure (per mmHg increase) | 0.206 (2.74) | 0.564 |
| Pulse pressure (per mmHg increase) | 0.894 (0.3) | 0.708 |
| Heart rate (per beat min<sup>-1</sup> increase) | 0.979 (0.02) | 0.368 |
| SuPAR (per ng/mL increase) | 1.112 (0.11) | 0.334 |
| hs-CRP (per mg/dL increase) | 0.883 (0.07) | 0.088 |
| Albumin (per g/dL increase) | 5.59 (1.3) | 0.185 |

hs-CRP, high-sensitivity C-reactive protein

Patient characteristics and disease progression after admission to the ICU are presented in Tables E3-E7 in the online data supplement. Vasoactive drugs were administered in both waves, but the doses of isoprenaline and norepinephrine were higher during the second wave (Tables E8, E9, and Figure E4 in the online data supplement). The incidence of complications after admission to the ICU was higher in the second wave, but not statistically significant (59% *vs.* 75%, p=0.3114) (see Table E10 in the online data supplement). We found no differences in acute coronary syndromes (23.5% vs. 9.3%, p=0.245) and ventilator-associated pneumonia (29.4% vs. 15.6%, p= 0.342) during ICU stay between patients of the first and second wave, respectively. The causes of death during the first wave were respiratory failure and sepsis (n=3). During the second wave, the most common causes of death were respiratory failure and multiple organ dysfunction syndrome (MODS) (n=49).

### Biomarkers of inflammation

In total, 66 (81.4%) patients had suPAR ≥ 5 ng/ml and 52 (64.2%) patients had suPAR ≥ 6 ng/ml. Mean suPAR was higher in the second wave, but after p-value adjustment, the difference between the two waves was statistically non-significant (7.33 *vs*. 9.3, p=0.023, adjusted p=0.1). We conducted a further analysis after dividing the patients into two subgroups, survivors *vs.* non-survivors, and found a positive correlation between suPAR and ICU length of stay in survivors (rho = 0.225, p=0.33). In addition, we found a statistically significant negative correlation between suPAR and ICU length of stay (rho= - 0.322, p=0.011) among non-survivors (Fig. 2).

**Figure 2.**
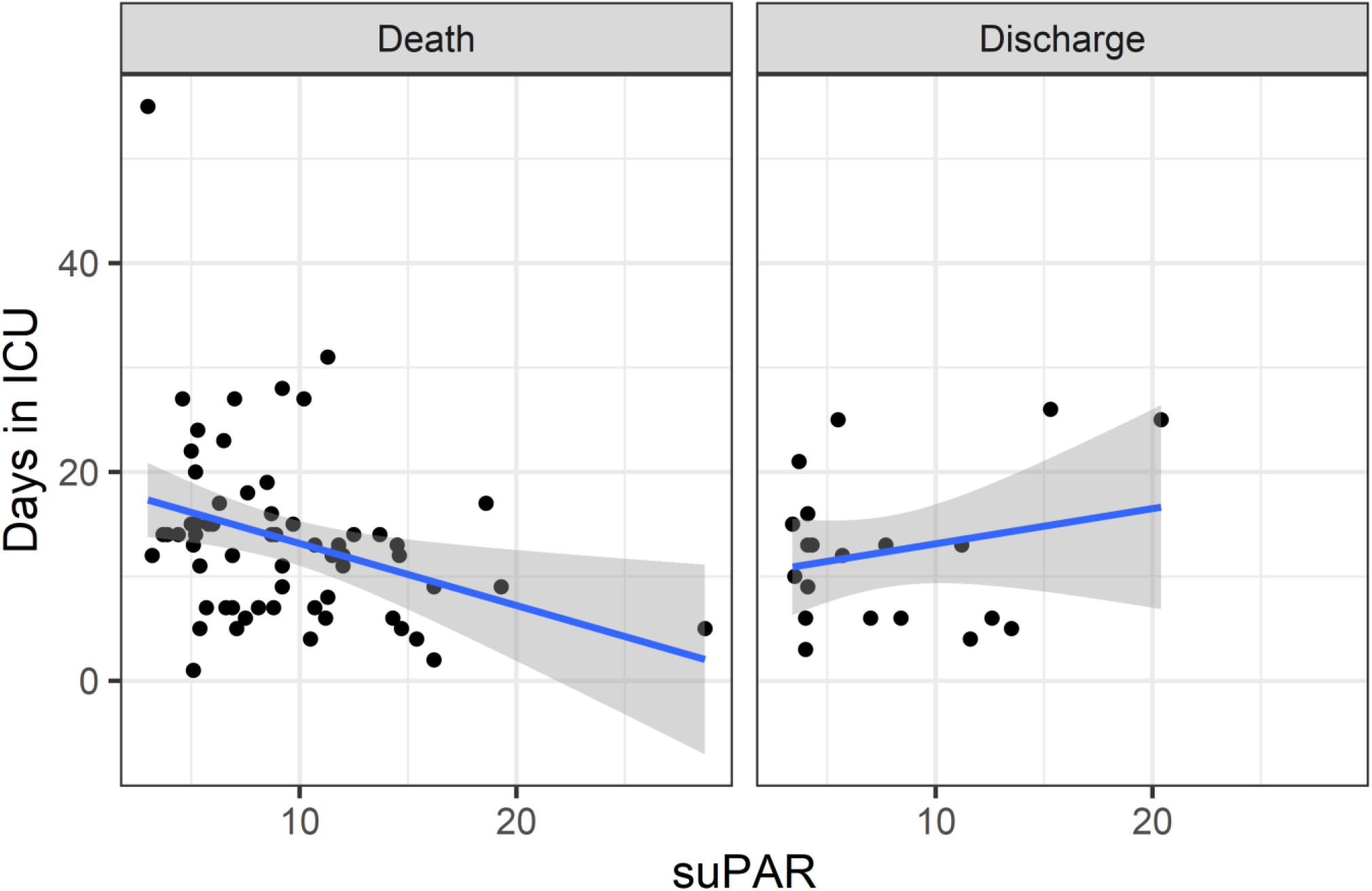
Correlation between suPAR and ICU length of stay in survivors (rho = 0.225, p=0.33) and non-survivors (rho= - 0.322, p=0.011).

Lower high-sensitivity C-reactive protein (hs-CRP) levels at presentation were associated with better survival (unadjusted p=0.034) (see Figure E1 in the online data supplement), but in multivariate analysis, hs-CRP was not associated with outcome. Ferritin at admission did not differ between the first and second wave, but lower ferritin levels were associated with prolonged days on ventilator and hospital stay (unadjusted p=0.033 and 0.026, respectively) (see Figures E2 and E3 in the online data supplement).

## Discussion

In this multicenter observational study, the most prominent finding was the difference in the severity of the disease and in overall survival between the first and second wave. Another significant characteristic of the second wave was the development of sinus bradycardia during hospitalization without clinically detectable myocardial necrosis nor being induced by drugs, indicating involvement of cardiac conduction system with SARS-CoV-2 infection that did not progress in parallel with pulmonary abnormalities (5).

The ICU mortality rate among patients with COVID-19 has been reported to be 30.6%, although it may increase up to 93% in mechanically ventilated patients with ARDS (11). In our study, the majority of critically ill patients was admitted during the second wave, with mortality being more than doubled (84.4% vs. 41.1%) despite the comparable patient characteristics between the two outbreak waves. This is the first report on mortality during the second wave in Greece, and is consistent with the upsurge in COVID-19 witnessed in Europe since September 2020 (12). Our findings are in contrast to studies from other countries reporting a lower disease severity and/or mortality in the second wave (13), but the degree of hypoxemia in our patients was among the most severe of those reported worldwide (1,4,7,8,13). Our results confirm that the risk of severe adverse outcomes and death in SARS-CoV-2 infected individuals shows extreme stratification according to age, which may improve understanding of the disease and patient management (14).

Other settings in our country have reported a lower mortality (32%) for overall ICU admissions during the first wave, highlighting the effects of an overburdened medical system on mortality rates rather than the severity of the disease *per se* (15). However, in a recently published Swedish registry-based cohort study of patients admitted from 6 March to 6 May 2020, the ICU mortality was even lower (23.1%) (16). The healthcare system in Sweden has never been overwhelmed, but until 10 November 2020 they reported one of the highest numbers of COVID-19 deaths per inhabitant globally (17). Therefore, the increased severity and mortality during the second wave in our study could be attributed to a more dangerous SARS-CoV-2 variant (18), which may have been introduced and transmitted regionally long before the first relevant announcement by the Hellenic National Organization for Public Health on December 23, 2020 (19). Continued community-based transmission of the European or other strains has been reported in several populations and may result in unknown mutations that are associated with severe disease, while they are usually identified much later from the peak of an outbreak (20). This is a major issue in countries like ours, which followed a strategy focused hard lockdowns and on hospital preparedness but failed to reinforce primary/community care and epidemiological surveillance (21,22).

A striking finding was the development of sinus bradycardia in 98.4% of the patients of the second wave without clinically detectable myocardial necrosis or after treatment with medications affecting heart rate. The average time for development of bradycardia was 5 and 10 days from admission and onset of symptoms, respectively (23), which is significantly shorter compared to other cohorts (5). The development of bradycardia in our patients may reflect the activation of the cholinergic nervous system in an effort to regulate the inflammatory response (24). However, recent evidence suggest that SARS-CoV-2 S1 protein may bind to nicotinic acetylcholine receptors and adversely affect their function by preventing acetylcholine’s action, causing dysregulation of the cholinergic anti-inflammatory pathway and leading to uncontrolled immune response and cytokine storm (24,25). Heart rate should normally have increased in these patients, but we did not observe such change. The combination of bradycardia and uncontrolled inflammatory response may be a novel clinical manifestation related to a new SARS-CoV-2 variant. In addition, the absence of myocardial necrosis together with the increase in D-Dimer in our patients may indicate an involvement through an uncharacterized pathway. In patients with COVID-19, dysfunctional endothelium may display a hypercoagulant/prothrombotic/pro-oxidant state and impairs microvascular reactivity (26). The latter, together with the increased levels of inflammatory mediators, enhances mechanical stress of cardiomyocytes and metabolic demands of conduction muscle cells, promoting metabolic instability and conduction disorders (23,27,28).

Although we did not find an association between bradycardia and outcome, our sample does not allow for a firm conclusion. Until now, patient characteristics, baseline ECG features, respiratory function, serum biomarkers of inflammation, and myocardial injury have an insufficient discriminatory power to identify subjects at increased risk for the development of new ECG changes (5). Nevertheless, these data suggest an inhibitory influence of the virus on cardiac conduction system and considering the high mortality in our study, we recommend close monitoring of patients with COVID-19 (28). Also, the use of drugs affecting the conduction system of the heart should be avoided in patients with sinus bradycardia or other conduction disorders, while the use of anticholinergics, such as atropine, could inhibit the protective effects of the cholinergic anti-inflammatory pathway.

A typical feature of our patients was the rapid progression to ARDS approximately 10 days after the onset of symptoms. However, time to intubation did not differ significantly between the two waves, although it was longer in the second one. The reasons for this were the concern that early intubation of every hypoxemic patient would be impossible due to the limited ICU capacity and resources, and the possible beneficial effects of avoiding early endotracheal intubation in significantly hypoxemic patients (29). Also, this practice was based in part on evidence from other ICUs showing that a strategy of early intubation was not associated with higher ICU-mortality, fewer ventilator-free days, or fewer ICU-free days than delayed or no intubation (15,30,31). Until now, optimal timing of initiation of invasive mechanical ventilation in COVID-19 patients with acute hypoxemic respiratory failure remains unknown.

In this cohort, overall survival and post-intubation complications did not differ significantly between rapid and delayed sequence intubation. Critically ill patients with COVID-19 usually have low functional residual capacity and minimal physiological reserve, which can prompt providers to try to secure the airway rapidly without adequate preoxygenation. However, rapid sequence intubation may result in immediate and profound desaturation even after a successful first intubation attempt (32). In patients with respiratory failure, apneic oxygenation has been recommended to prevent desaturation during intubation (33,34), but this technique did not prevent desaturation during or after intubation in our patients (7,8,35). We agree that patients with COVID-19 require detailed planning and strategy for tracheal intubation and therefore, delayed sequence intubation may be a valuable technique for maximizing safety, especially in ward patients with limited staff and equipment availability (8,32). This technique can offer an alternative to rapid sequence intubation in patients who will not tolerate preoxygenation or other peri-intubation procedures. However, the superiority of one technique vs. the other must be investigated in randomized controlled studies. In our study, ICU and hospital length of stay were higher during the first wave. This is attributed mainly to the severity of COVID-19 and the increased mortality during the second wave, and less to the differences in complications between the two waves. For example, the percentage of ICU patients who were diagnosed with ventilator-associated pneumonia in our study is lower when compared to other studies (36,37). Considering that the mean number of days on ventilator was higher in the first wave, the most possible explanation for the higher (but not statistically significant) prevalence of ventilator-associated pneumonia during the second wave may be the increased predisposition due to the increased severity of lung damage caused by COVID-19 (38). Also, the cumulative risk of secondary sepsis increases with ICU stay (38), and bacterial DNA and toxins have been discovered in all severely ill patients with COVID-19 (39,40). Until now, clinically relevant sepsis and septic shock have been reported in up to 60% of cases, which is in accordance with our results (first wave 47%, second wave 50%) (41,42). On the other hand, respiratory failure and MODS were key determinants of survival during the second wave. MODS was related to the direct and indirect pathogenic features of SARS-CoV-2 and was induced by the hyperinflammation and humoral and cell-mediated immune response (42). However, hs-CRP was not an independent predictor of disease severity in our study.

suPAR has been shown to provide important indications for required early admission and treatment in non-COVID-19 patients; the TRIAGE III trial including 4420 patients reported that suPAR ranged between 2.6 and 4.7 ng/ml in 30-day survivors and between 6.7 and 11.8 ng/ml in 30-day non-survivors (43). The dysregulation of the urokinase plasminogen activator/urokinase plasminogen activator receptor system may be also a main cause of organ failure in patients with SARS-CoV-2 infection and especially in those with chronic inflammation (44). A suPAR level ≥ 6 ng/ml has been independently associated with the development of severe respiratory failure in patients with COVID-19 (45–47), which is in agreement with our results. Moreover, we observed for the first time that a higher suPAR at presentation was correlated with earlier death after ICU admission among non-survivors, while a lower suPAR (but still >6 ng/ml) was correlated with prolonged ICU length of stay in critically ill patients who survived. This finding is important because ferritin levels were markedly increased in our patients, but their difference between the two waves did not reach statistical significance. Although ferritin has been repeatedly reported as a potent marker of disease severity (48), a smaller increase in ferritin levels at admission was associated with prolonged days on ventilator and hospital stay, possibly reflecting a reduced immune activation due to the milder form of the disease at that point. Considering that patients with COVID-19 have heterogeneous cytokine profiles, suPAR seems extremely promising as a prognostic marker in those with severe disease and after ICU admission (49,50). Two ongoing large multicenter observational studies whose primary purpose is to characterize levels of suPAR among various biomarkers of inflammation and its association with in-hospital outcomes of patients with COVID-19 are expected to conclude by the end of spring 2021.

The study has several strengths. It is a multicenter study that relied on collection of clinical, laboratory, and outcome data throughout the COVID-19 hospitalization during two successive outbreak waves, capturing a diverse patient population. Data collection was systematic and all patients admitted/intubated during the period April 1^st^ to December 31^st^ were enrolled. Our sample was limited to patients consecutively hospitalized specifically for COVID-19 and without receiving any specific treatment besides dexamethasone, allowing for a better description of the effect of SARS-CoV-2 infection on different organs of the human body. The major limitations of the present study are the relatively small sample and its observational nature. Despite the careful analysis, it is not possible to fully account for all potential confounders and therefore, the study cannot be trusted *per se* as a basis of clinical decision. However, our findings have significant implications for a better understanding of the epidemiology of COVID-19 and for better planning and organization in the future.

The present study adds to the growing body of research that indicates differences in the severity of COVID-19 and in overall survival between successive outbreak waves. Moreover, it provides additional evidence with respect to the involvement of cardiac conduction system with SARS-CoV-2. These findings may help in the classification of novel phenotypes for informing treatment strategy and for identifying populations that may benefit from early admission to ICU.

## Data Availability

Data can be made available upon request through a collaborative process

## Acknowledgements

Nothing to acknowledge

