## supplemental file for "Clinical course and outcomes of critically ill COVID-19 patients in two successive pandemic waves"

### SUPPLEMENTARY MATERIAL

#### FIGURES

**Figure E1**

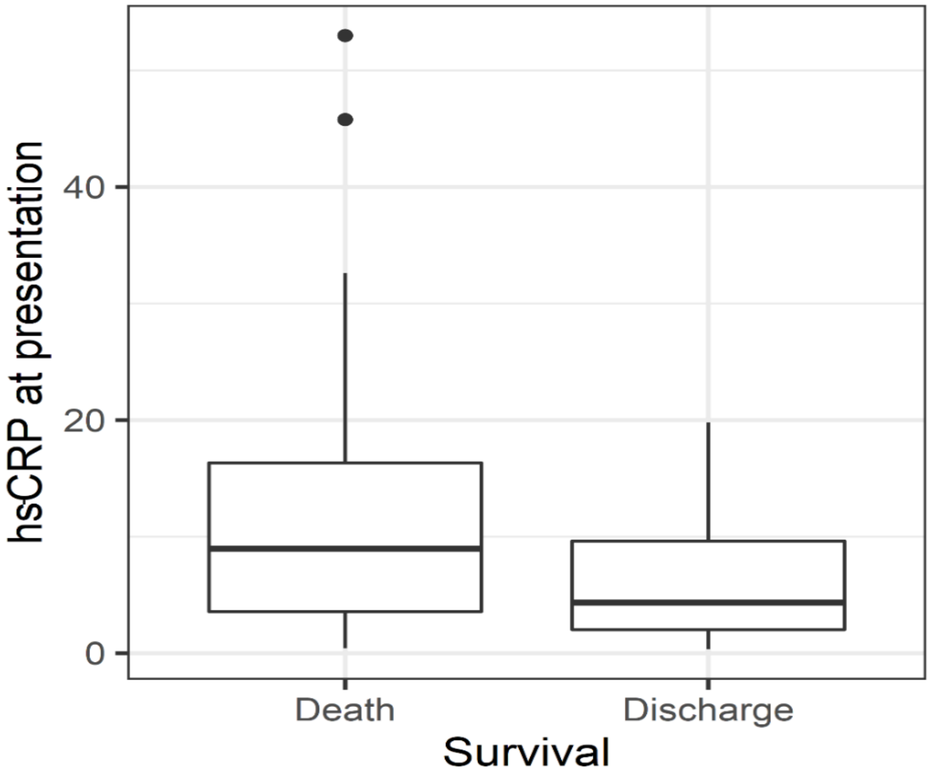

**Figure E2**

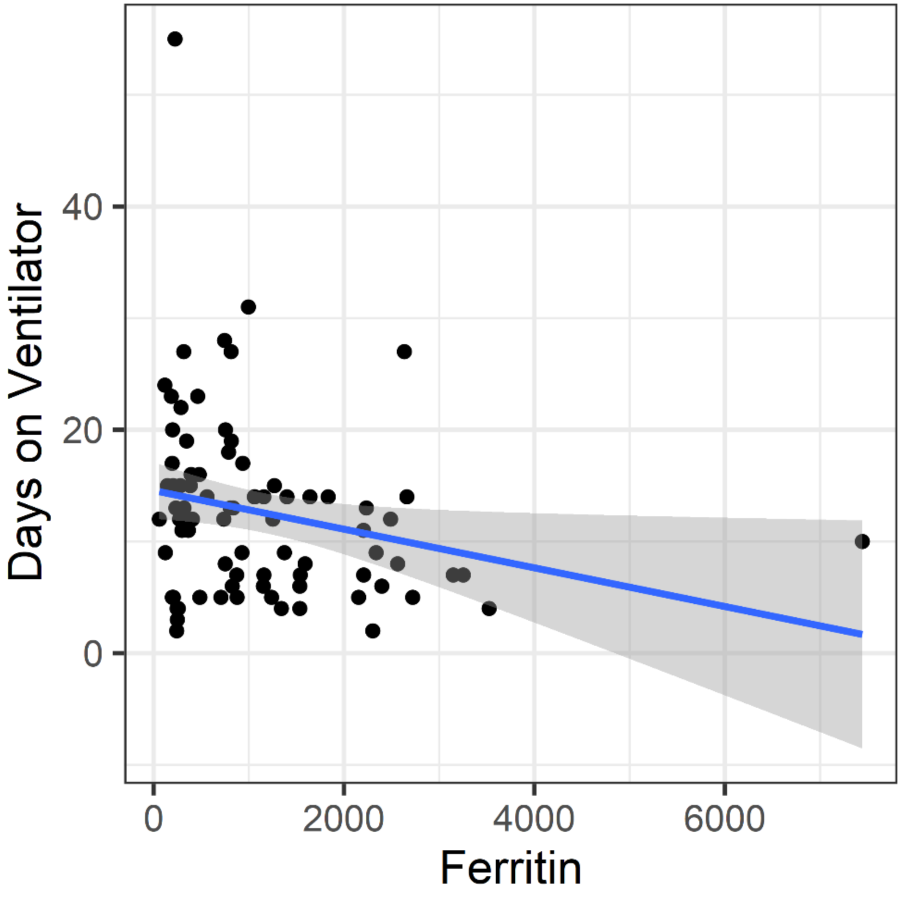

**Figure E3**

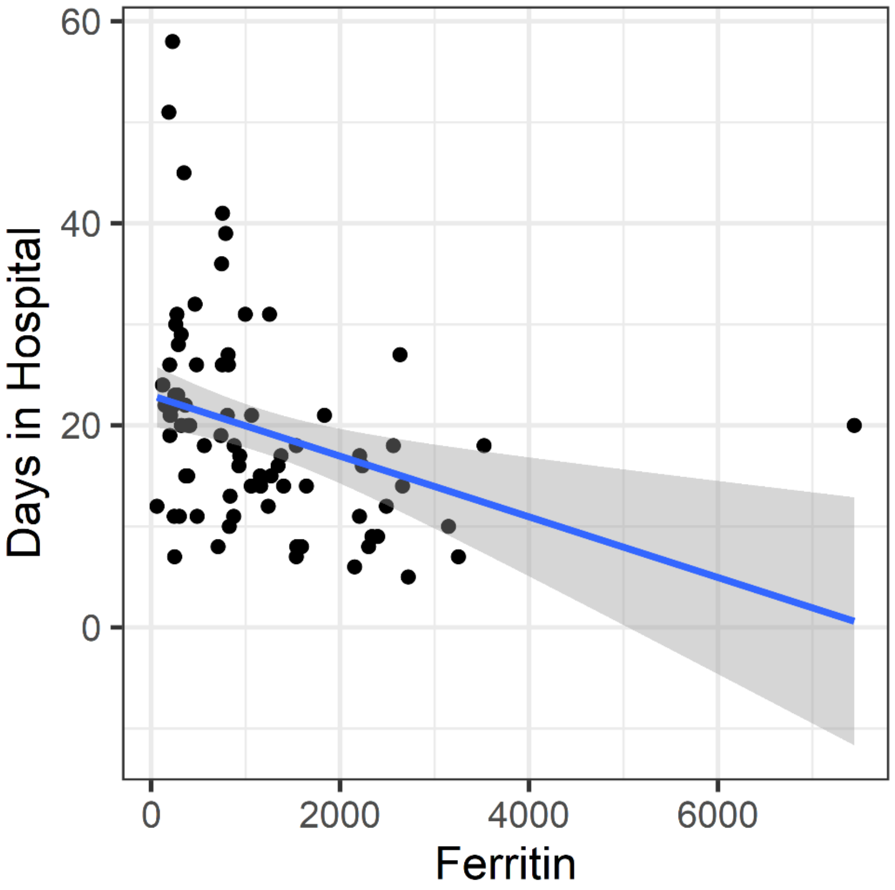

**Figure E4**

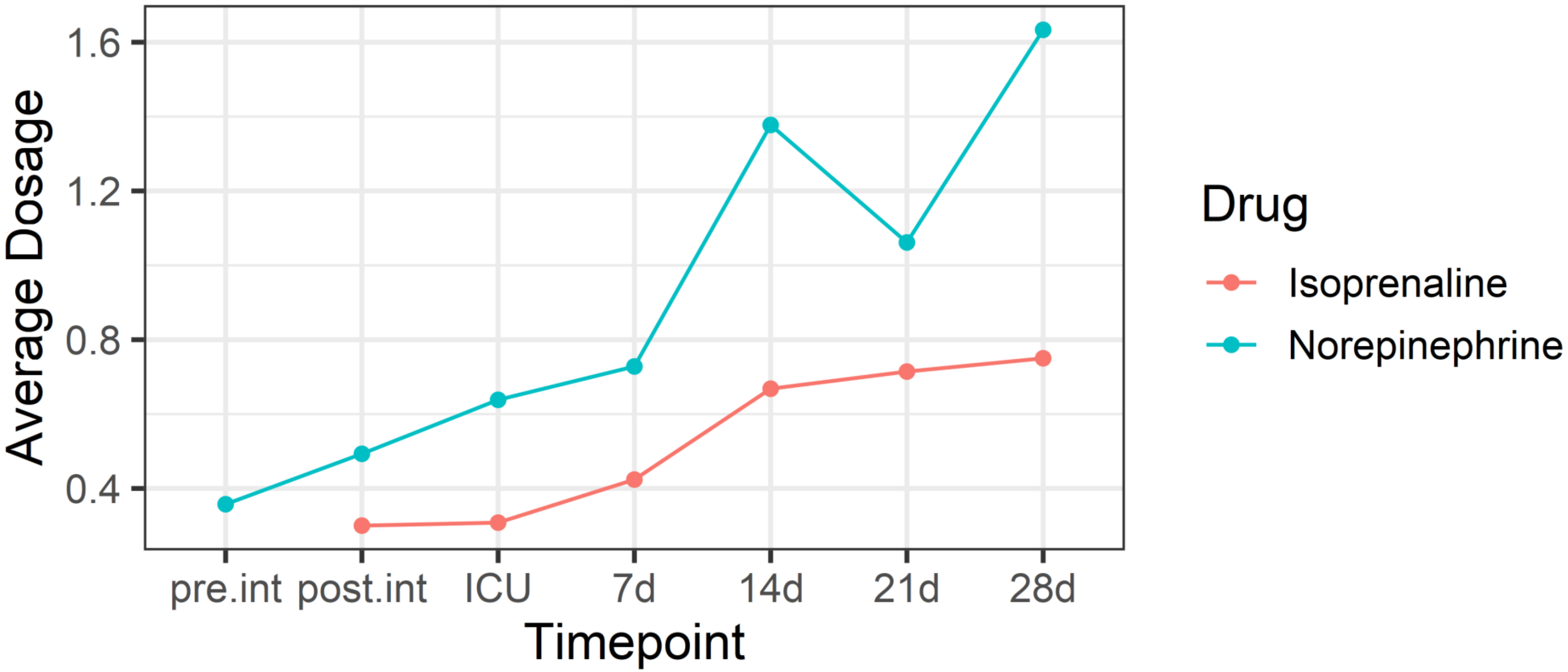


In both waves, the doses of isoprenaline and norepinephrine increased during hospitalization (p<0.001)

pre.int, pre-intubation; post.int, post-intubation; ICU, admission to the Intensive Care Unit; d, days

### TABLES

Table E1. Pre-intubation characteristics of the patients

| | | First wave (n=17) | Second wave (n=64) | Adjusted p-value |
|---|---|---|---|---|
| Decision to intubate | Hours after admission (h), mean (95% CI) | 95 (53.8 - 136.2) | 106.2 (80 - 132.3) | 0.861 |
| Emergency degree | Intubation required in less than 1 hour, n (%) | 10 (59%) | 54 (84%) | 0.259 |
| | Intubation required in more than 1 hour, n (%) | 3 (5%) | 4 (6%) | |
| | Intubation required without any delay, n (%) | 4 (6%) | 6 (9%) | |
| Intubation site | Ward, n (%) | 17 (100%) | 56 (88%) | 0.747 |
| | Emergency Department, n (%) | 0 (0%) | 5 (8%) | |
| | Intensive Care Unit, n (%) | 0 (0%) | 3 (5%) | |
| Breathing help before intubation | High-flow nasal cannula, n (%) | 17 (100%) | 45 (70%) | 0.25 |
| | Facemask with $O_2$ reservoir, n (%) | 0 (0%) | 14 (22%) | |
| CPAP | Yes, n (%) | 0 (0%) | 7 (11%) | 0.74 |
| | No, n (%) | 17 (100%) | 57 (89%) | |
| Bradycardia before intubation | Sinus - absolute, n (%) | 13 (76%) | 50 (78%) | 0.83 |

| | | | | | |
|---|---|---|---|---|---|
| | Sinus - absolute, First-degree AV block, n (%) | | 0 (0%) | 1 (2%) | |
| | Sinus - relative, n (%) | | 14 (22%) | 13 (20%) | |
| | Symptomatic | Yes, n (%) | 0 (0%) | 3 (5%) | |
| | | No, n (%) | 17 (100%) | 61 (95%) | |
| Need of new/modification of vasopressor/inotrope support before intubation | Norepinephrine, n (%) | | 1 (6%) | 19 (30%) | 0.28 |
| | No, n (%) | | 16 (94%) | 45 (70%) | |
| Anticipated difficult airway management? | Beard, n (%) | | 1 (6%) | 5 (8%) | 0.91 |
| | Neck stiffness, n (%) | | 1 (6%) | 2 (3%) | |
| | Neck stiffness, Obesity, n (%) | | 0 (0%) | 1 (2%) | |
| | Obesity, n (%) | | 4 (24%) | 10 (16%) | |
| | Reduced mouth opening (< 3 cm), n (%) | | 1 (6%) | 1 (2%) | |
| | Reduced mouth opening (< 3 cm), Obesity, n (%) | | 1 (6%) | 0 (0%) | |
| | Reduced thyro-mental distance, n (%) | | 1 (6%) | 3 (5%) | |
| | Reduced thyro-mental distance, Obesity, n (%) | | 0 (0%) | 1 (2%) | |
| | Retrognathia, n (%) | | 1 (6%) | 1 (2%) | |
| | No, n (%) | | 7 (41%) | 38 (59%) | |
| | Head-elevation (30-45°), n (%) | | 3 (18%) | 17 (27%) | 0.77 |

| | | | | |
|---|---|---|---|---|
| Patient position during pre-oxygenation | Ramp, n (%) | 5 (30%) | 11 (17%) | |
| | Supine, n (%) | 9 (53%) | 36 (56%) | |
| Device used for pre-oxygenation | High-flow nasal cannula, n (%) | 16 (94%) | 61 (95%) | 1 |
| | Standard nasal cannula, n (%) | 1 (6%) | 3 (5%) | |
| Oxygen administration during laryngoscopy | High-flow nasal cannula, n (%) | 0 (0%) | 45 (70%) | **<0.001** |
| | Standard nasal cannula, n (%) | 4 (24%) | 9 (14%) | |
| | No, n (%) | 13 (76%) | 10 (16%) | |
| Induction | Delayed sequence intubation, n (%) | 7 (41%) | 26 (41%) | 1 |
| | Rapid sequence intubation, n (%) | 10 (59%) | 38 (59%) | |
| | Midazolam, n (%) | 0 (0%) | 12 (19%) | |
| | Fentanyl, n (%) | 17 (100%) | 64 (100%) | |
| | Ketamine, n (%) | 0 (0%) | 12 (19%) | |
| | Propofol, n (%) | 14 (82%) | 54 (84%) | |
| | Etomidate, n (%) | 17 (100%) | 52 (81%) | |
| | Lidocaine, n (%) | 16 (94%) | 42 (66%) | |
| | Rocuronium, n (%) | 17 (100%) | 60 (94%) | |
| | Succinylcholine, n (%) | 0 (0%) | 4 (6%) | |

| | | | | |
|---|---|---|---|---|
| Sellick maneuver applied | No, n (%) | 11 (65%) | 51 (80%) | 0.75 |
| | Yes, n (%) | 6 (35%) | 13 (20%) | |
| Laryngoscopy | Direct with Macintosh blade, n (%) | 14 (82%) | 47 (73%) | 0.76 |
| | Direct with Macintosh blade, Videolaryngoscopy, n (%) | 0 (0%) | 5 (8%) | |
| | Videolaryngoscopy, n (%) | 3 (18%) | 12 (19%) | |
| First pass success | No, n (%) | 4 (24%) | 7 (11%) | 0.74 |
| | Yes, n (%) | 13 (76%) | 57 (89%) | |
| Difficult intubation | No, n (%) | 13 (76%) | 57 (89%) | 0.74 |
| | Yes, n (%) | 4 (24%) | 7 (11%) | |
| APACHE II score, mean (95% CI) | | 11.11 (10.44 - 11.79) | 10.71 (10.31 - 11.13) | 0.463 |
| SOFA score, mean (95% CI) | | 5.35 (3.74 - 6.97) | 4.25 (3.99 - 4.51) | 0.861 |

CPAP, continuous positive airway pressure; AV, atrioventricular

Table E2. Peri-intubation hemodynamic and metabolic parameters [mean (95% CI)]

| | **First wave (n=17)** | **Second wave (n=64)** | **Adjusted p-value** |
|---|---|---|---|
| *Prior to intubation* | | | |
| Systolic arterial pressure (mmHg) | 119.52 (111.62 - 127.44) | 126.93 (122.42 - 131.46) | 0.276 |
| Diastolic arterial pressure (mmHg) | 60.35 (56.09 - 64.61) | 66.37 (63.12 - 69.63) | 0.209 |
| Mean arterial pressure (mmHg) | 80.13 (75.64 - 84.63) | 86.45 (83.15 - 89.75) | 0.135 |
| Pulse pressure (mmHg) | 59.17 (51.57 - 66.79) | 60.46 (56.73 - 64.2) | 0.861 |
| Heart rate ($min^{-1}$) | 61.17 (54.99 - 67.37) | 60.7 (57.65 - 63.76) | 0.813 |
| Respiratory rate ($min^{-1}$) | 34.88 (31.45 - 38.32) | 34.03 (32.75 - 35.31) | 0.86 |
| $SpO_2$ (%) | 78.35 (75.16 - 81.55) | 74.57 (72.28 - 76.88) | 0.278 |
| Capillary refill time (s) | 3.52 (3.08 - 3.98) | 3.09 (2.76 - 3.43) | 0.281 |
| Temperature (°C) | 37.95 (37.57 - 38.35) | 38.52 (38.35 - 38.7) | 0.052 |
| P/F Ratio | 54.88 (53.08 - 56.68) | 52.15 (50.19 - 54.12) | 0.234 |
| pH | 7.46 (7.45 - 7.48) | 7.44 (7.43 - 7.45) | 0.159 |
| $PaO_2$ (mmHg) | 54.88 (53.08 - 56.68) | 52.15 (50.19 - 54.12) | 0.234 |
| $PaCO_2$ (mmHg) | 39.47 (37.38 - 41.56) | 37.07 (36.09 - 38.07) | 0.171 |
| $Na^+$ (mmol/L) | 139.31 (136.9 - 141.73) | 137.67 (136.49 - 138.86) | 0.457 |

| | | | |
|---|---|---|---|
| $K^+$ (mmol/L) | 3.9 (3.65 - 4.16) | 3.99 (3.86 - 4.14) | 0.831 |
| $Ca^{++}$ (mmol/L) | 8.19 (7.99 - 8.4) | 8.16 (8.04 - 8.29) | 0.831 |
| Glu (mg/dL) | 133 (111.12 - 154.89) | 151.34 (137.09 - 165.6) | 0.455 |
| $HCO_3^-$ (mmol/L) | 27.82 (26.26 - 29.39) | 28.03 (27.09 - 28.97) | 0.859 |
| BE (mmol/L) | 2.52 (1.83 - 3.22) | 2.52 (2.18 - 2.87) | 0.954 |
| Lactate (mmol/L) | 1.54 (1.3 - 1.78) | 1.56 (1.41 - 1.72) | 1 |
| Glasgow Coma Scale/Score | 14.64 (14.34 - 14.96) | 14.75 (14.6 - 14.9) | 0.488 |
| Urine output (ml/Kg/h) | 0.48 (0.46 - 0.51) | 0.54 (0.5 - 0.58) | 0.448 |
| *Immediate post-intubation period (mechanical ventilation)* | | | |
| Systolic arterial pressure (mmHg) | 98.82 (91.72 - 105.93) | 101.93 (97.5 - 106.38) | 0.601 |
| Diastolic arterial pressure (mmHg) | 51.7 (46.83 - 56.59) | 55.39 (52.21 - 58.58) | 0.457 |
| Mean arterial pressure (mmHg) | 67.41 (62.27 - 72.55) | 70.91 (67.47 - 74.35) | 0.499 |
| Pulse pressure (mmHg) | 47.11 (41.79 - 52.45) | 46.56 (43.95 - 49.17) | 0.831 |
| Heart rate ($min^{-1}$) | 76.05 (72.02 - 80.09) | 57.78 (54.84 - 60.73) | **<0.001** |
| Respiratory rate ($min^{-1}$) | 25 (23.11 - 26.89) | 27.01 (26.28 - 27.75) | 0.065 |
| $SpO_2$ (%) | 94.47 (93.44 - 95.5) | 94.82 (94.48 - 95.17) | 0.459 |
| Capillary refill time (s) | 4.76 (4.27 - 5.26) | 3.85 (3.43 - 4.29) | 0.114 |

| | | | |
|---|---|---|---|
| Temperature (°C) | 37.95 (37.57 - 38.35) | 38.52 (38.35 - 38.7 ) | 0.052 |
| P/F Ratio | 129.29 (107.08 - 151.51) | 95.95 (86.43 - 105.48 ) | **0.045** |
| pH | 7.41 (7.4 - 7.43) | 7.39 (7.38 - 7.4) | 0.083 |
| $PaO_2$ (mmHg) | 129.29 (107.08 - 151.51) | 95.95 (86.43 - 105.48) | **0.045** |
| $PaCO_2$ (mmHg) | 40 (37.83 - 42.17) | 37.92 (36.89 - 38.96) | 0.21 |
| $Na^+$ (mmol/L) | 139.85 (137.46 - 142.25) | 139.98 (138.83 - 141.14) | 0.886 |
| $K^+$ (mmol/L) | 4.21 (3.9 - 4.53) | 4.25 (4.11 - 4.39) | 0.861 |
| $Ca^{++}$ (mmol/L) | 8.03 (7.78 - 8.28) | 8.01 (7.88 - 8.16) | 0.898 |
| Glu (mg/dL) | 155.1 (124.13 - 186.07) | 160.7 (147.17 - 174.23) | 0.822 |
| $HCO_3^-$ (mmol/L) | 26.29 (25.05 - 27.54) | 24.98 (24.07 - 25.9) | 0.347 |
| BE (mmol/L) | 2.34 (1.63 - 3.05) | 2.07 (1.72 - 2.42) | 0.649 |
| Lactate (mmol/L) | 1.61 (1.35 - 1.87) | 1.64 (1.49 - 1.8) | 1 |

$SpO_2$, peripheral capillary oxygen saturation; $PaO_2$, arterial partial pressure of oxygen; $PaCO_2$, arterial partial pressure of carbon dioxide

Table E3. Patient parameters in the Intensive Care Unit at 48h [mean (95% CI)]

| | **First wave (n=17)** | **Second wave (n=64)** | **Adjusted p-value** |
|---|---|---|---|
| APACHE II score | 12.88 (10.99 - 14.77) | 14.57 (13.82 - 15.34) | 0.124 |
| SOFA score | 10.76 (9.58 - 11.95) | 11.84 (11.31 - 12.38) | 0.288 |
| Systolic arterial pressure (mmHg) | 129.94 (124.91 - 134.97) | 126.5 (123.67 - 129.33) | 0.561 |
| Diastolic arterial pressure (mmHg) | 59.64 (51.22 - 68.08) | 65.35 (63.19 - 67.53) | 0.474 |
| Mean arterial pressure (mmHg) | 87.47 (82.25 - 92.69) | 84.51 (81.53 - 87.5) | 0.677 |
| Pulse pressure (mmHg) | 68.29 (62.5 - 74.09) | 61.12 (57.66 - 64.59) | 0.195 |
| Heart rate ($min^{-1}$) | 75.88 (74.23 - 77.54) | 55.82 (53.4 - 58.26) | **<0.001** |
| Respiratory rate ($min^{-1}$) | 25.17 (23.69 - 26.67) | 26.98 (26.43 - 27.53) | 0.101 |
| $SpO_2$ (%) | 93.7 (93.2 - 94.21) | 92.1 (91.46 - 92.76) | 0.081 |
| Capillary refill time (s) | 3.88 (3.48 - 4.28) | 3.96 (3.75 - 4.19) | 0.859 |
| Temperature (°C) | 38.36 (37.98 - 38.75) | 38.59 (38.44 - 38.76) | 0.36 |
| White blood cells (K/uL) | 9.52 (8.46 - 10.57) | 9.8 (8.57 - 11.04) | 0.557 |
| Hemoglobin (g/dL) | 12.25 (11.6 - 12.9) | 11.58 (11.08 - 12.08) | 0.356 |
| Platelets (K/uL) | 249.88 (204.78 - 294.99) | 242.92 (214.61 - 271.23) | 0.822 |
| BUN (mg/dL) | 55.06 (35.42 - 74.71) | 79.9 (67.47 - 92.34) | 0.101 |

| | | | |
|---|---|---|---|
| Creatinine (mg/dL) | 1.14 (0.79 - 1.5) | 1.47 (1.15 - 1.79) | 0.775 |
| Protein (g/dl) | 5.4 (5.12 - 5.7) | 5.41 (5.23 - 5.6) | 0.822 |
| Albumin (g/dl) | 3.16 (2.97 - 3.35) | 2.9 (2.73 - 3.08) | 0.052 |
| hs-CRP (mg/dL) | 3.23 (0.85 - 5.62) | 9.73 (7.89 - 11.57) | **0.001** |
| Ferritin (ng/mL) | 3926.82 (1367.2 - 6486.45) | 1646.66 (1290.54 - 2002.79) | 0.346 |
| D-Dimer (ng/mL) | 1370.29 (504.73 - 2235.85) | 1493.77 (1116.56 - 1871) | 0.198 |
| Lactate dehydrogenase (IU/L) | 689.82 (242.51 - 1137.14) | 575.56 (493.78 - 657.35) | 0.406 |
| P/F Ratio | 165.52 (137.15 - 193.91) | 146.46 (130.16 - 162.78) | 0.272 |
| pH | 7.39 (7.38 - 7.42) | 7.33 (7.33 - 7.35) | **<0.001** |
| $PaO_2$ (mmHg) | 113.41 (94.03 - 132.81) | 104.17 (95.43 - 112.93) | 0.463 |
| $PaCO_2$ (mmHg) | 40.23 (38.32 - 42.16) | 47.57 (45.67 - 49.48) | **0.002** |
| $Na^+$ (mmol/L) | 144.02 (141.43 - 146.62) | 142.05 (141.26 - 142.85) | 0.278 |
| $K^+$ (mmol/L) | 4.59 (4.35 - 4.84) | 4.28 (4.18 - 4.4) | 0.112 |
| $Ca^{++}$ (mmol/L) | 7.69 (7.45 - 7.93) | 7.88 (7.75 - 8.02) | 0.278 |
| Glu (mg/dL) | 158.93 (131.98 - 185.89) | 152.37 (143.81 - 160.94 ) | 0.898 |
| $HCO_3^-$ (mmol/L) | 25.82 (24.61 - 27.03) | 19.92 (18.76 - 21.09) | **<0.001** |
| BE (mmol/L) | 1.06 (0.33 - 1.8) | 0.18 (-0.53 - 0.89) | 0.719 |

| | | | |
|---|---|---|---|
| Lactate (mmol/L) | 1.66 (1.42 - 1.91) | 2.25 (1.98 - 2.53) | 0.115 |

$SpO_2$, peripheral capillary oxygen saturation; BUN, Blood urea nitrogen; hs-CRP, high-sensitivity C-reactive protein; $PaO_2$, arterial partial pressure of oxygen; $PaCO_2$, arterial partial pressure of carbon dioxide

Table E4. Patient parameters in the Intensive Care Unit at 7 days [mean (95% CI)]

| | **First wave (n=17)** | **Second wave (n=64)** | **Adjusted p-value** |
|---|---|---|---|
| APACHE II score | 12.64 (10.27 - 15.02) | 17.1 (15.78 - 18.44) | **0.027** |
| SOFA score | 11.47 (10.36 - 12.58) | 12.81 (12.19 - 13.43) | 0.069 |
| Systolic arterial pressure (mmHg) | 126.76 (123.05 - 130.48) | 121.84 (119.02 - 124.67) | 0.278 |
| Diastolic arterial pressure (mmHg) | 62.64 (59.02 - 66.28) | 61.56 (59.49 - 63.63) | 0.358 |
| Mean arterial pressure (mmHg) | 84.01 (81.31 - 86.73) | 81.74 (79.89 - 83.59) | 0.34 |
| Pulse pressure (mmHg) | 64.11 (58.9 - 69.34) | 60.43 (57.3 - 63.57) | 0.358 |
| Heart rate ($min^{-1}$) | 71.17 (67.62 - 74.73) | 56.07 (53.88 - 58.27) | **<0.001** |
| Respiratory rate ($min^{-1}$) | 24.05 (22.61 - 25.51) | 27.26 (26.45 - 28.08) | **0.002** |
| $SpO_2$ (%) | 91.88 (90.17 - 93.6) | 89.84 (89.2 - 90.49) | **0.016** |
| Capillary refill time (s) | 4 (3.55 - 4.45) | 4.29 (4.01 - 4.58) | 0.561 |
| Temperature (°C) | 38.17 (37.84 - 38.51) | 38.31 (38.14 - 38.5) | 0.642 |
| White blood cells (K/uL) | 9.46 (6.79 - 12.14) | 10.47 (9 – 11.78) | 0.997 |
| Hemoglobin (g/dL) | 11.47 (10.52 - 12.43) | 12.38 (8.87 - 15.89) | 0.262 |
| Platelets (K/uL) | 245.82 (193.82 - 297.83) | 206.09 (175.83 – 236.35) | 0.273 |
| BUN (mg/dL) | 69.48 (43.3 - 95.68) | 103.26 (88.15 - 118.37) | 0.055 |

| | | | |
|---|---|---|---|
| Creatinine (mg/dL) | 1.19 (0.54 - 1.85) | 1.52 (1.22 - 1.84) | 0.156 |
| Protein (g/dl) | 5.5 (4.92 - 6.1) | 5.29 (5.07 - 5.53) | 1 |
| Albumin (g/dl) | 2.98 (2.76 - 3.22) | 2.78 (2.6 - 2.97) | 0.13 |
| hs-CRP (mg/L) | 2.34 (0.15 - 4.53) | 11.74 (9.56 - 13.92) | **<0.001** |
| Ferritin (ng/mL) | 6523.14 (-2682.93 - 15729.22) | 3031.83 (1515.54 - 4548.13) | 0.833 |
| D-Dimer (ng/mL) | 1181.17 (420.99 - 1941.36) | 1300.59 (1030.28 - 1570.91) | 0.159 |
| Lactate dehydrogenase (IU/L) | 991.64 (-13.93 - 1997.22) | 578.89 (499.42 - 658.36) | 0.278 |
| IL-6 (pg/ml) | 2121.73 (-4080.24 - 8323.71) | 27.32* | 0.669 |
| $FiO_2$ (%) | 63.05 (55.09 - 71.03) | 77.35 (72.61 - 82.11) | **0.045** |
| P/F Ratio | 164.29 (132.06 - 196.52) | 134.5 (116.46 - 152.54) | 0.137 |
| pH | 7.36 (7.34 - 7.38) | 7.26 (7.24 - 7.28) | **<0.001** |
| $PaO_2$ (mmHg) | 101.62 (87.18 - 116.07) | 91.36 (84.24 - 98.48) | 0.358 |
| $PaCO_2$ (mmHg) | 46.7 (43.5 - 49.92) | 52.57 (50.36 - 54.8) | 0.059 |
| $Na^+$ (mmol/L) | 142.62 (139.89 - 145.15) | 142.68 (141.28 - 144.08) | 0.813 |
| $K^+$ (mmol/L) | 4.51 (4.2 - 4.84) | 4.28 (4.15 - 4.41) | 0.36 |
| $Ca^{++}$ (mmol/L) | 7.66 (7.27 - 8.07) | 7.65 (7.51 - 7.81) | 0.969 |
| Glu (mg/dL) | 143.58 (121.94 - 165.24) | 138.06 (128.64 - 147.48) | 0.74 |

| | | | |
|---|---|---|---|
| $HCO_3^-$ (mmol/L) | 23.82 (21.94 - 25.71) | 15.91 (14.75 - 17.08) | **<0.001** |
| BE (mmol/L) | -0.16 (-0.99 - 0.68) | -5.15 (-6.19 - -4.12) | **<0.001** |
| Lactate (mmol/L) | 1.89 (1.5 - 2.29) | 5.36 (4.49 - 6.23) | **<0.001** |

* No other values to calculate 95% CI. There was a shortage of IL-6 reagents during the 2nd wave.

$SpO_2$, peripheral capillary oxygen saturation; BUN, Blood urea nitrogen; hs-CRP, high-sensitivity C-reactive protein; IL-6, interleukin-6; $FiO_2$, Fraction of inspired oxygen; $PaO_2$, arterial partial pressure of oxygen; $PaCO_2$, arterial partial pressure of carbon dioxide

Table E5. Patient parameters in the Intensive Care Unit at 14 days [mean (95% CI)]

| | **First wave (n=17)** | **Second wave (n=31)** | **Adjusted p-value** |
|---|---|---|---|
| APACHE II score | 11.82 (8.76 - 14.88) | 17.45 (15.49 - 19.41) | 0.053 |
| SOFA score | 12.26 (10.95 - 13.58) | 12.61 (11.69 - 13.53) | 0.74 |
| Systolic arterial pressure (mmHg) | 120.76 (112.08 - 129.45) | 116.29 (108.65 - 123.93) | 0.682 |
| Diastolic arterial pressure (mmHg) | 65.7 (58.3 - 73.11) | 64.32 (58.49 - 70.15) | 0.898 |
| Mean arterial pressure (mmHg) | 84.05 (76.44 - 91.68) | 81.68 (75.4 - 87.97) | 0.829 |
| Pulse pressure (mmHg) | 55.05 (50.97 - 59.15) | 52 (48.47 - 55.53) | 0.545 |
| Heart rate ($min^{-1}$) | 69.05 (64.86 - 73.26) | 58.45 (55.41 - 61.49) | **0.002** |
| Respiratory rate ($min^{-1}$) | 23.52 (20.74 - 26.32) | 27.06 (25.73 - 28.4) | 0.179 |
| $SpO_2$ (%) | 91.23 (88.81 - 93.66) | 87.41 (86.1 - 88.74) | 0.086 |
| Capillary refill time (s) | 4.23 (3.54 - 4.93) | 5.71 (5.14 - 6.28) | **0.018** |
| Temperature (°C) | 37.82 (37.56 - 38.09) | 38.27 (37.59 - 38.97) | 0.474 |
| White blood cells (K/uL) | 11.08 (7.96 - 14.22) | 475.3 (-202.58 - 1153.18) | 0.476 |
| Hemoglobin (g/dL) | 10.1 (9.45 - 10.77) | 1441.25 (-1482.17 - 4364.69) | 0.682 |
| Platelets (K/uL) | 182.94 (132.55 - 233.33) | 8829.13 (-3559.02 - 21217.28) | 0.719 |
| BUN (mg/dL) | 70.84 (46.59 - 95.11) | 87.95 (72.12 - 103.79) | 0.305 |

| | | | |
|---|---|---|---|
| Creatinine (mg/dL) | 1.53 (0.79 - 2.29) | 1.46 (1.05 - 1.89) | 0.822 |
| Protein (g/dl) | 5.36 (4.9 - 5.83) | 5.09 (4.74 - 5.44) | 0.75 |
| Albumin (g/dl) | 3.05 (2.83 - 3.29) | 2.83 (2.48 - 3.19) | 0.145 |
| hs-CRP (mg/L) | 3.57 (0.92 - 6.23) | 22.15 (4.04 - 40.26) | **0.001** |
| Ferritin (ng/mL) | 6284.71 (-3378.47 - 15947.9) | 2609.4 (1639.12 - 3579.69) | 0.358 |
| D-Dimer (ng/mL) | 1281.11 (510.5 - 2051.74) | 1055.38 (796.17 - 1314.6) | 0.346 |
| Lactate Dehydrogenase (IU/L) | 731.23 (-31.41 - 1493.88) | 560.54 (386.98 - 734.12) | 0.441 |
| IL-6 (pg/ml) | 344.02 (-181.02 - 869.07) | 27.32* | 0.561 |
| $FiO_2$ (%) | 59.05 (50 - 68.12) | 68.83 (63.07 - 74.6) | 0.201 |
| P/F Ratio | 185.35 (151.67 - 219.04) | 164.29 (130.45 - 198.13) | 0.278 |
| pH | 7.34 (7.29 - 7.39) | 242.66 (-238.11 - 723.44) | 0.053 |
| $PaO_2$ (mmHg) | 100.6 (91.22 - 110) | 99.06 (87.32 - 110.81) | 0.499 |
| $PaCO_2$ (mmHg) | 46 (43.04 - 48.96) | 49.42 (47.11 - 51.73) | 0.195 |
| $Na^+$ (mmol/L) | 201.87 (77.81 - 325.93) | 141.97 (139.81 - 144.13) | 0.463 |
| $K^+$ (mmol/L) | 4.16 (3.84 - 4.49) | 4.37 (4.21 - 4.54) | 0.42 |
| $Ca^{++}$ (mmol/L) | 7.68 (7.21 - 8.15) | 7.91 (7.66 - 8.16) | 0.736 |
| Glu (mg/dL) | 137.67 (115.74 - 159.61) | 145.93 (132.73 - 159.14) | 0.358 |

| | | | |
|---|---|---|---|
| $HCO_3^-$ (mmol/L) | 20.97 (17.33 - 24.61) | 15.26 (13.33 - 17.2) | 0.105 |
| BE (mmol/L) | -1.86 (-3.65 - -0.08) | -5.67 (-7.2 - -4.15) | **0.045** |
| Lactate (mmol/L) | 3.09 (1.91 - 4.27) | 6.31 (4.88 - 7.76) | **0.034** |

* No other values to calculate 95% CI. There was a shortage of IL-6 reagents during the 2nd wave.

$SpO_2$, peripheral capillary oxygen saturation; BUN, Blood urea nitrogen; hs-CRP, high-sensitivity C-reactive protein; IL-6, interleukin-6; $FiO_2$, Fraction of inspired oxygen; $PaO_2$, arterial partial pressure of oxygen; $PaCO_2$, arterial partial pressure of carbon dioxide

Table E6. Patient parameters in the Intensive Care Unit at 21 days [mean (95% CI)]

| | **First wave (n=6)** | **Second wave (n=9)** | **Adjusted p-value** |
|---|---|---|---|
| APACHE II score | 8.83 (4.56 - 13.1) | 19.44 (17.01 - 21.88) | **0.031** |
| SOFA score | 9.33 (7.17 - 11.5) | 12.44 (10.95 - 13.94) | 0.083 |
| Systolic arterial pressure (mmHg) | 127.33 (105.95 - 148.71) | 115.55 (104.88 - 126.23) | 0.463 |
| Diastolic arterial pressure (mmHg) | 71.5 (60.78 - 82.22) | 64 (54.45 - 73.55) | 0.463 |
| Mean arterial pressure (mmHg) | 90.11 (76.72 - 103.5) | 81.18 (71.85 - 90.52) | 0.358 |
| Pulse pressure (mmHg) | 55.83 (40.87 - 70.8) | 54.55 (44.09 - 65.02) | 1 |
| Heart rate ($min^{-1}$) | 73.66 (62.31 - 85.02) | 57.55 (53.83 - 61.28) | 0.101 |
| Respiratory rate ($min^{-1}$) | 22 (15.88 - 28.12) | 27.55 (25.38 - 29.73) | 0.241 |
| $SpO_2$ (%) | 93.16 (88.74 - 97.59) | 87.44 (86.05 - 88.84) | 0.101 |
| Capillary refill time (s) | 3.83 (2.15 - 5.51) | 5.66 (5 - 6.33) | 0.107 |
| Temperature (°C) | 36.93 (36.24 - 37.63) | 37.85 (37.63 - 38.08) | 0.092 |
| White blood cells (K/uL) | 7.36 (3.76 - 10.98) | 1052.55 (-1353.6 - 3458.71) | 0.488 |
| Hemoglobin (g/dL) | 9.43 (7.4 - 11.47) | 9.17 (8.31 - 10.05) | 0.859 |
| Platelets (K/uL) | 203.5 (121.46 - 285.54) | 59419.11 (-77090.88 - 195929.1) | 0.755 |
| BUN (mg/dL) | 62.2 (-1.13 - 125.53) | 53.45 (29.58 - 77.33) | 0.86 |

| | | | |
|---|---|---|---|
| Creatinine (mg/dL) | 0.78 (0.58 - 0.98) | 0.62 (0.39 - 0.85) | 0.481 |
| Protein (g/dl) | 5.28 (4.24 - 6.32) | 5.09 (4.58 - 5.6) | 0.822 |
| Albumin (g/dl) | 3.01 (2.34 - 3.68) | 2.41 (1.98 - 2.84) | 0.294 |
| hs-CRP (mg/L) | 2.47 (-2.5 - 7.44) | 8.47 (2.13 - 14.82) | 0.111 |
| Ferritin (ng/mL) | 719.7 (-18.15 - 1457.55) | 2199.76 (-784.3 - 5183.82) | 0.406 |
| D-Dimer (ng/mL) | 535.5 (301.95 - 769.05) | 754.87 (590.97 - 918.78) | 0.234 |
| Lactate Dehydrogenase (IU/L) | 298.66 (164.36 - 432.97) | 502.77 (113.3 - 892.26) | 0.755 |
| $FiO_2$ (%) | 50.66 (29.43 - 71.9) | 74.88 (64.35 - 85.42) | 0.14 |
| P/F Ratio | 235.5 (174.18 - 296.82) | 109.55 (88.57 - 130.54) | **0.028** |
| pH | 7.35 (7.25 - 7.46) | 7.24 (7.2 – 7.29) | 0.19 |
| $PaO_2$ (mmHg) | 109.55 (100.83 - 118.28) | 79.01 (70.16 - 87.86) | **0.022** |
| $PaCO_2$ (mmHg) | 42.5 (37 - 48) | 47.44 (45.68 - 49.21) | 0.262 |
| $Na^+$ (mmol/L) | 145.9 (137.82 - 153.98) | 145.66 (141.33 - 150) | 0.859 |
| $K^+$ (mmol/L) | 4.17 (3.85 - 4.51) | 4.05 (3.77 - 4.34) | 0.612 |
| $Ca^{++}$ (mmol/L) | 8.29 (7.83 - 8.75) | 7.38 (6.75 - 8.03) | 0.117 |
| Glu (mg/dL) | 159.98 (108.71 - 211.26) | 132.11 (87.59 - 176.63) | 0.488 |
| $HCO_3^-$ (mmol/L) | 22.33 (15.01 - 29.65) | 13.74 (11.08 - 16.41) | 0.189 |

| | | | |
|---|---|---|---|
| BE (mmol/L) | -1.35 (-7.64 - 4.94) | -7.2 (-9.55 - -4.85) | 0.201 |
| Lactate (mmol/L) | 2.65 (-0.53 - 5.83) | 7.68 (5.52 - 9.85) | 0.066 |

$SpO_2$, peripheral capillary oxygen saturation; BUN, Blood urea nitrogen; hs-CRP, high-sensitivity C-reactive protein; $FiO_2$, Fraction of inspired oxygen; $PaO_2$, arterial partial pressure of oxygen; $PaCO_2$, arterial partial pressure of carbon dioxide

Table E7. Patient parameters in the Intensive Care Unit at 28 days [mean (95% CI)]

| | **First wave (n=5)** | **Second wave (n=4)** | **Adjusted p-value** |
|---|---|---|---|
| APACHE II score | 12.25 (-3.41 - 27.91) | 21 (16.32 - 25.68) | 0.499 |
| SOFA score | 9 (3.96 - 14.04) | 11.75 (9.36 - 14.14) | 0.474 |
| Systolic arterial pressure (mmHg) | 117 (87.03 - 146.97) | 100.75 (97.47 - 104.03) | 0.356 |
| Diastolic arterial pressure (mmHg) | 64.8 (42.36 - 87.24) | 51 (43.54 - 58.46) | 0.356 |
| Mean arterial pressure (mmHg) | 82.2 (57.37 - 107.03) | 67.58 (62.14 - 73.03) | 0.358 |
| Pulse pressure (mmHg) | 52.2 (43.06 - 61.34) | 49.75 (42.71 - 56.79) | 0.7 |
| Heart rate ($min^{-1}$) | 74 (63.46 - 84.54) | 59.75 (57.36 - 62.14) | 0.21 |
| Respiratory rate ($min^{-1}$) | 23.2 (13.29 - 33.11) | 31.25 (27.27 - 35.23) | 0.391 |
| $SpO_2$ (%) | 90.2 (80.81 - 99.59) | 83.5 (82.58 - 84.42) | 0.356 |
| Capillary refill time (s) | 5.2 (0.36 - 10.04) | 7.75 (5.75 - 9.75) | 0.355 |
| Temperature (°C) | 37.34 (36.02 - 38.66) | 37.95 (36.86 - 39.04) | 0.555 |
| White blood cells (K/uL) | 8.68 (6 - 11.36) | 11.07 (4.57 - 17.58) | 0.463 |
| Hemoglobin (g/dL) | 9.44 (6.28 - 12.6) | 8.4 (7.8 - 9) | 0.833 |
| Platelets (K/uL) | 227.8 (126.64 - 328.96) | 413.5 (268.17 - 558.83) | 0.086 |
| BUN (mg/dL) | 54.52 (5.45 - 103.59) | 57.27 (12.72 - 101.83) | 0.833 |

| | | | |
|---|---|---|---|
| Creatinine (mg/dL) | 0.91 (0.5 - 1.34) | 0.54 (0.08 - 1.01) | 0.358 |
| Protein (g/dL) | 5.14 (3.18 - 7.11) | 5.26 (4.57 - 5.97) | 0.833 |
| Albumin (g/dL) | 2.54 (1.43 - 3.65) | 2.72 (2.01 - 3.43) | 0.94 |
| hs-CRP (mg/L) | 3.48 (-1.51 - 8.47) | 5.98 (2.31 - 9.66) | 0.555 |
| Ferritin (ng/mL) | 3765.98 (-4836.01 - 12367.97) | 1622.73 (-1062.02 - 4307.49) | 0.735 |
| D-Dimer (ng/mL) | 641.2 (165 - 1117.4) | 817.33 (20.08 - 1614.59) | 0.735 |
| Lactate Dehydrogenase (IU/L) | 638.8 (-482.68 - 1760.28) | 430.75 (192.23 - 669.27) | 0.488 |
| $FiO_2$ (%) | 50.6 (20.43 - 80.77) | 85.25 (67.37 - 103.13) | 0.278 |
| P/F Ratio | 238.2 (117.27 - 359.13) | 91.5 (48.99 - 134.01) | 0.278 |
| pH | 7.32 (7.12 - 7.52) | 7.21 (7.1 - 7.33) | 0.358 |
| $PaO_2$ (mmHg) | 101.8 (81.36 - 122.26) | 75.89 (50.15 - 101.63) | 0.278 |
| $PaCO_2$ (mmHg) | 143.6 (-138.58 - 425.78) | 51.5 (45.62 - 57.38) | 0.455 |
| $Na^+$ (mmol/L) | 142.88 (133.56 - 152.2) | 147.57 (146.2 - 148.95) | 0.576 |
| $K^+$ (mmol/L) | 4.51 (2.73 - 6.3) | 4.42 (3.73 - 5.12) | 0.358 |
| $Ca^{++}$ (mmol/L) | 8.25 (7.67 - 8.84) | 7.67 (7.1 - 8.24) | 0.278 |
| Glu (mg/dL) | 118.12 (99.06 - 137.18) | 127.25 (69.49 - 185.01) | 1 |
| $HCO_3^-$ (mmol/L) | 21.2 (11.21 - 31.19) | 41.85 (-56.98 - 140.68) | 0.831 |

| | | | |
|---|---|---|---|
| BE (mmol/L) | -2.74 (-11.64 - 6.16) | -10.17 (-13.64 - -6.71) | 0.358 |
| Lactate (mmol/L) | 4.32 (-3.48 - 12.12) | 10.75 (6.01 - 15.49) | 0.356 |

$SpO_2$, peripheral capillary oxygen saturation; BUN, Blood urea nitrogen; hs-CRP, high-sensitivity C-reactive protein; $FiO_2$, Fraction of inspired oxygen; $PaO_2$, arterial partial pressure of oxygen; $PaCO_2$, arterial partial pressure of carbon dioxide

Table E8. Number of patients requiring vasopressors/inotropes and average doses

| | **Patients - First wave, n (%)** | **Patients - Second wave, n (%)** | **Dose - First wave[a]** | **Dose - Second wave[a]** | **p-value[b]** |
|---|---|---|---|---|---|
| Norepinephrine | 17 (100%) | 64 (100%) | 0.54 (0.54) mcg/kg/min | 0.82 (0.82) mcg/kg/min | 0.051 |
| Isoprenaline | 2 (11.7%) | 43 (67.1%) | 1.03 (0.19) mcg/kg/min | 0.42 (0.24) mcg/kg/min | 0.026 |
| Vasopressin | 1 (3.03%) | 33 (51.6%) | 0.06 (NA) units/min[c] | 0.21 (0.86) units/min | -[c] |
| Dobutamine | 0 (0%) | 3 (4.69%) | -[c] | 1.2 (0.9) mcg/kg/min | -[c] |

[a] Data presented as mean (standard deviation)

[b] Doses between the two waves

[c] More than two samples need to compute

NA, non-applicable

Table E9. Dose of vasopressors/inotropes at specific time points

| | Peri-intubation period | | After admission to the Intensive Care Unit | | | | |
|---|---|---|---|---|---|---|---|
| | **Pre-intubation**[a] | **Post-intubation**[a] | **48 h**[a] | **7 days**[a] | **14 days**[a] | **21 days**[a] | **28 days**[a] |
| Norepinephrine, mcg/kg/min | 0.36 (0.28) | 0.49 (0.54) | 0.64 (0.6) | 0.73 (0.62) | 1.38 (3) | 1.06 (0.68) | 1.63 (0.5) |
| Isoprenaline, mcg/kg/min | NA[b] | 0.3 (NA)[b] | 0.31 (0.31) | 0.42 (0.34) | 0.67 (0.32) | 0.71 (0.3) | 0.75 (0.41) |
| Vasopressin, units/min | NA[b] | NA[b] | 0.89 (1.85) | 0.31 (1.14) | 0.05 (0.01) | 0.05 (0.01) | 0.06 (NA)[b] |
| Dobutamine, mcg/kg/min | NA[b] | NA[b] | NA[b] | NA[b] | 2.1 (NA)[b] | 0.2 (NA)[b] | 0.8 (0.57) |

[a] Data presented as mean (standard deviation)

[b] More than two samples need to compute

NA, non-applicable

Table E10. Complications after admission to the Intensive Care Unit

| **At 48 hours** | | **First wave (n=17)** | **Second wave (n=64)** | **Adjusted p-value** |
|---|---|---|---|---|
| Bradycardia | No, n (%) | 17 (100%) | 2 (3%) | **<0.001** |
| | Sinus - absolute, n (%) | 0 (0%) | 52 (81%) | |
| | Sinus - relative, n (%) | 0 (0%) | 10 (16%) | |
| Sepsis, n (%) | | 0 (0%) | 1 (2%) | 0.997 |
| MODS, n (%) | | 0 (0%) | 1 (2%) | |
| Acute kidney injury, n (%) | | 0 (0%) | 0 (0%) | 1 |
| No complications, n (%) | | 17 (100%) | 62 (97%) | 0.997 |
| **At day 7** | | **First wave (n=17)** | **Second wave (n=64)** | **Adjusted p-value** |
| Bradycardia | No, n (%) | 17 (100%) | 10 (16%) | **0.001** |
| | Sinus - absolute, n (%) | 0 (0%) | 50 (78%) | |
| | Sinus - relative, n (%) | 0 (0%) | 4 (6%) | |
| VAP, n (%) | | 2 (12%) | 4 (%) | 1 |
| VAP, Sepsis, n (%) | | 1 (6%) | 3 (5%) | |
| Sepsis, n (%) | | 3 (18%) | 14 (22%) | |
| MODS, n (%) | | 0 0(%) | 1 (2%) | |

| | | | | |
|---|---|---|---|---|
| MODS, VAP, n (%) | | 0 (0%) | 1 (2%) | |
| Sepsis, Acute coronary syndrome, n (%) | | 0 (0%) | 1 (2%) | |
| Sepsis, MODS, n (%) | | 0 (0%) | 2 (3%) | |
| Stroke, n (%) | | 0 (0%) | 1 (2%) | |
| Stroke, MODS, n (%) | | 0 (0%) | 1 (2%) | |
| Acute kidney injury, n (%) | | 2 (12%) | 1 (2%) | 0.643 |
| No complications, n (%) | | 11 (65%) | 36 (56%) | 1 |
| **At day 14** | | **First wave (n=17)** | **Second wave (n=31)** | **Adjusted p-value** |
| Bradycardia | No, n (%) | 16 (94%) | 3 (10%) | **<0.001** |
| | Sinus - absolute, n (%) | 1 (6%) | 25 (81%) | |
| | Sinus - relative, n (%) | 0 (0%) | 3 (10%) | |
| VAP, n (%) | | 2 (12%) | 2 (6%) | 0.895 |
| VAP, Sepsis, n (%) | | 2 (12%) | 1 (3%) | |
| Sepsis, n (%) | | 3 (18%) | 10 (%) | |
| Sepsis, Acute coronary syndrome, n (%) | | 1 (6%) | 1 (3%) | |
| Sepsis, Arrythmia, n (%) | | 1 (6%) | 0 (0%) | |
| Sepsis, MODS, n (%) | | 1 (6%) | 3 (10%) | |

| | | | | |
|---|---|---|---|---|
| Acute coronary syndrome, n (%) | | 0 (0%) | 2 (6%) | |
| Acute coronary syndrome, Arrythmia, MODS, n (%) | | 0 (0%) | 1 (3%) | |
| Acute coronary syndrome, Heart failure, MODS, n (%) | | 0 (0%) | 1 (3%) | |
| Acute coronary syndrome, MODS, n (%) | | 0 (0%) | 2 (6%) | |
| MODS, VAP, Sepsis, n (%) | | 0 (0%) | 1 (3%) | |
| MODS, VAP, Acute coronary syndrome, n (%) | | 0 (0%) | 1 (3%) | |
| Sepsis, Arrythmia, Stroke, n (%) | | 0 (0%) | 1 (3%) | |
| Acute kidney injury, n (%) | | 1 (6%) | 3 (10%) | 0.426 |
| No complications, n (%) | | 7 (41%) | 5 (16%) | 0.895 |
| **At day 21** | | **First wave (n=6)** | **Second wave (n=9)** | **Adjusted p-value** |
| Bradycardia | No, n (%) | 6 (100%) | 0 (0%) | **0.012** |
| | Sinus - relative, n (%) | 0 (0%) | 1 (11%) | |
| | Sinus - absolute, n (%) | 0 (0%) | 8 (89%) | |
| Sepsis, n (%) | | 1 (17%) | 6 (67%) | 0.278 |
| Sepsis, MODS, n (%) | | 1 (17%) | 0 (0%) | |
| MODS, n (%) | | 0 (0%) | 1 (11%) | |
| VAP, Sepsis, n (%) | | 0 (0%) | 1 (11%) | |

| | | | | |
|---|---|---|---|---|
| Acute kidney injury, n (%) | | 2 (33%) | 2 (22%) | 1 |
| No complications, n (%) | | 4 (67%) | 1 (11%) | 0.278 |
| **At day 28** | | **First wave (n=5)** | **Second wave (n=4)** | **Adjusted p-value** |
| Bradycardia | No, n (%) | 4 (80%) | 0 (0%) | 0.051 |
| | Sinus - relative, n (%) | 0 (0%) | 1 (25%) | |
| | Sinus - absolute, n (%) | 1 (20%) | 3 (75%) | |
| Sepsis, n (%) | | 1 (20%) | 1 (25%) | 1 |
| MODS, Sepsis, n (%) | | 1 (20%) | 0 (0%) | - |
| MODS, n (%) | | 0 (0%) | 1 (25%) | - |
| VAP, Sepsis, n (%) | | 0 (0%) | 1 (25%) | - |
| Sepsis, n (%) | | 0 (0%) | 1 (25%) | - |
| Sepsis, Heart failure, n (%) | | 0 (0%) | 1 (25%) | - |
| Acute kidney injury, n (%) | | 0 (0%) | 0 (0%) | 1 |
| No complications, n (%) | | 3 (60%) | 0 (0%) | - |

MODS, Multiple Organ Dysfunction Syndrome, VAP, ventilator-associated pneumonia
